# Viral cultures, Polymerase Chain Reaction Cycle Threshold Values and Viral Load Estimation for SARS-CoV-2 Infectious Potential Assessment in Hematopoietic Stem Cell and Solid Organ Transplant Patients: A Systematic Review

**DOI:** 10.1101/2022.03.01.22271684

**Authors:** Tom Jefferson, Elizabeth A. Spencer, John M. Conly, Elena C. Rosca, Susanna Maltoni, Jon Brassey, Igho J. Onakpoya, David H. Evans, Carl J. Heneghan, Annette Plüddemann

## Abstract

**Background:** Organ transplant recipients are at increased vulnerability to SARS-CoV-2 due to immunosuppression and may pose a continued transmission risk especially within hospital settings. Detailed case reports including symptoms, viral load and infectiousness, defined by the presence of replication-competent viruses in culture, provide an opportunity to examine the relationship between clinical course, burden and contagiousness, and provide guidance on release from isolation.

**Objectives:** We performed a systematic review to investigate the relationship in transplant recipients between serial SARS-CoV-2 RT-PCR cycle threshold (Ct) value or cycle of quantification value (Cq), or other measures of viral burden and the likelihood and duration of the presence of infectious virus based on viral culture including the influence of age, sex, underlying pathologies, degree of immunosuppression, and/or vaccination on this relationship.

**Methods:** We searched LitCovid, medRxiv, Google Scholar and WHO Covid-19 databases, from 1 November 2019 until 31 December 2021. We included studies reporting relevant data for transplantees with SARS-CoV-2 infection: results from serial RT-PCR testing and viral culture data from the same respiratory samples. We assessed methodological quality using five criteria, and synthesised the data narratively and graphically.

**Results:** We included 10 case reports and case series reporting on 38 transplantees. We observed a relationship between proxies of viral burden and likelihood of shedding replication-competent SARS-CoV-2. Two individuals shed replication-competent viruses over 100 days after infection onset. Lack of standardisation of testing and reporting platforms precludes establishing a definitive viral burden cut-off. However, most transplantees stopped shedding competent viruses when the RT-PCR cycle threshold was above 30 despite differences across platforms.

**Conclusions:** Viral burden is a reasonable proxy for infectivity when considered within the context of the clinical status of each patient. Standardised study design and reporting are essential to standardise guidance based on an increasing evidence base.

## Introduction

Hematopoietic stem cell transplant (HSCT) and solid organ transplant (SOT) recipients have significant immunosuppression, affecting both cellular and humoral immunity, and less favourable outcomes with Severe Acute Respiratory Virus Syndrome 2 (SARS-CoV-2) infection, due to the immunosuppression and/or to pre-existing comorbidities^1^. Immunosuppression associated with transplantation places patients at risk for prolonged carriage and shedding of several respiratory viruses^2^. However, identification of respiratory viral shedding, recently by reverse transcriptase polymerase chain reaction (RT-PCR), depending on the testing platform does not correlate with the presence of replication-competent virus^3^. Accordingly, we sought to perform a systematic review of RT-PCR testing and viral culture of SARS-CoV-2, focussing on people receiving solid organ or hematopoietic stem cell transplants, following our published protocol^4^.

Our research questions were:

1. What is the correlation between serial SARS-CoV-2 RT-PCR cycle threshold (Ct) value or cycle of quantification value (Cq), or other measures of viral burden and the likelihood of producing replication-competent virus?
2. What is the likelihood and duration of the presence of infectious virus based on viral culture, among transplant recipients with SARS-CoV-2 infection?
3. What is the influence of age, sex, underlying pathologies and degree of immunosuppression on infectiousness of SARS-CoV-2?
4. What is the relationship of vaccination status on infectiousness with SARS-CoV-2?

We included studies reporting serial Cts from sequential RT-PCR testing or other measures of viral burden such as RNA gene copies of respiratory samples (from nasopharyngeal or throat specimens) along with viral culture data on the same samples, from patients about to receive a transplant or who were post-transplant, with SARS-CoV-2 infection.

## Methods

### Search Strategy

We searched the following electronic databases: LitCovid, medRxiv, Google Scholar and the WHO Covid-19 database from November 2019 until December 31, 2021. No language restrictions were applied.

The literature search terms were: (coronavirus OR covid-19 OR SARS-CoV-2) AND (immunosuppressed OR immunocompromised OR transplant OR immunosuppression OR “immune deficient” OR HIV) AND (CPE OR “cytopathic effect” OR “Viral culture” OR “virus culture” OR vero OR “virus replication” OR “viral replication” OR “cell culture” or “viral load” OR “viral threshold” OR “log copies” OR “cycle threshold”).

### Screening

Four reviewers independently screened titles and abstracts to identify studies for consideration of full text. Full text screening was performed in duplicate and disagreements arbitrated by a fifth reviewer.

### Inclusion criteria

We included studies reporting serial Cts from sequential RT-PCR testing, or RNA gene copies of respiratory samples (nasopharyngeal, throat, sputum, bronchoalveolar lavage, endotracheal tube secretions) AND viral culture data from the same samples from patients about to receive a transplant or post-transplant with SARS-CoV-2 infection. We included primary studies provided they reported sufficient information to extract quantitative data on the PCR testing and the viral culture for each included individual. Studies that included transplant and non-transplant patients were included if we could ascertain the results separately. Studies reported only in poster or abstract form were excluded. Reviews were excluded but the reference lists screened for potential relevant primary studies.

### Exclusion criteria

We excluded studies using post-mortem samples only and non-respiratory samples only. We did not include studies of non-transplant patients or those not attempting viral cultures.

### Data extraction

One reviewer extracted data, which was independently checked by a second reviewer. Disagreements were arbitrated by a third reviewer. Data were extracted on study type and study characteristics, including population, setting, sampling and laboratory methods, clinical information, prescribed treatments, vaccination status, laboratory findings, and clinical outcomes. For three studies we sought clarification from the corresponding authors.

### Quality assessment

We assessed the quality of included studies according to five criteria:

1. Were the criteria for diagnosing a case clearly reported and appropriate?
2. Was the reporting of patient/population characteristics including clinical symptoms, treatments with degree of immunosuppression and outcomes adequate?
3. Was the study period, including follow-up, sufficient to adequately assess any potential relationship between viral burden measures and likelihood of producing replication-competent virus and the rise in neutralising antibodies? We defined sufficient as more than one observation.
4. Were the methods used to obtain RT-PCR results replicable, generalisable and appropriate? We considered that each study should establish the relationship between their Ct values and the target gene copy number, using internal standards.
5. Were the methods used to obtain viral culture results replicable and appropriate? We considered the methods used should, at a minimum, include a description of specimen sampling and management, preparation, media and cell line used, exclusion of contamination or co-infection (use of good controls and appropriate antibacterials and antimycotics and possible use of gene sequencing if available), and results of inspection of culture.

### Data reporting and pooling

We reported study flow according to PRISMA reporting standards^5^. We reported study characteristics including age, sex, clinical symptoms, treatments and events in the participants in tabular form. We presented data on disease burden measures and on viral culture in tabular form. For studies reporting more than one patient participant, data were extracted related to each participant if available. We plotted median, interquartile ranges (IQRs) and outliers for viral culture results in relation to the duration of symptoms, and individual study plots to day 120 of viral culture results and cycle thresholds.

We were unable to meta-analyse the data on PCR cycle counts/RNA log copies and viral culture, due to a lack of detailed information on laboratory practices, assays and because of the absence of internal controls in some studies, and heterogeneous sampling. We therefore reviewed the studies narratively, and where possible presented the results graphically within the limitations noted.

We analysed the relationship between cycle threshold, days of onset of symptoms and likelihood of shedding replication-competent virus by presenting the data on a scatter plot

## Results

The literature search identified 12,988 titles for screening. Of these 63 underwent full-text review. A total of 53 studies were excluded after full-text analysis: reasons are reported in the list of excluded studies (see Supplementary File: List of excluded studies)

The 10 included studies (Figure 1 reported data for 30 transplant patients (7 females and 23 males): renal (21), cardiac (5), bone marrow transplant (9), liver (2), bilateral lung (1). For one study the number of males and females was not available for the relevant patient group. ^6^ The 38 patients were in seven countries: Saudi Arabia^7^, France ^8 9^, Germany ^10 11 12^, Austria ^13^, Denmark ^14^, Canada ^15^ and the USA^6^, and were aged between 26 and 77 years old.

**Figure 1.**
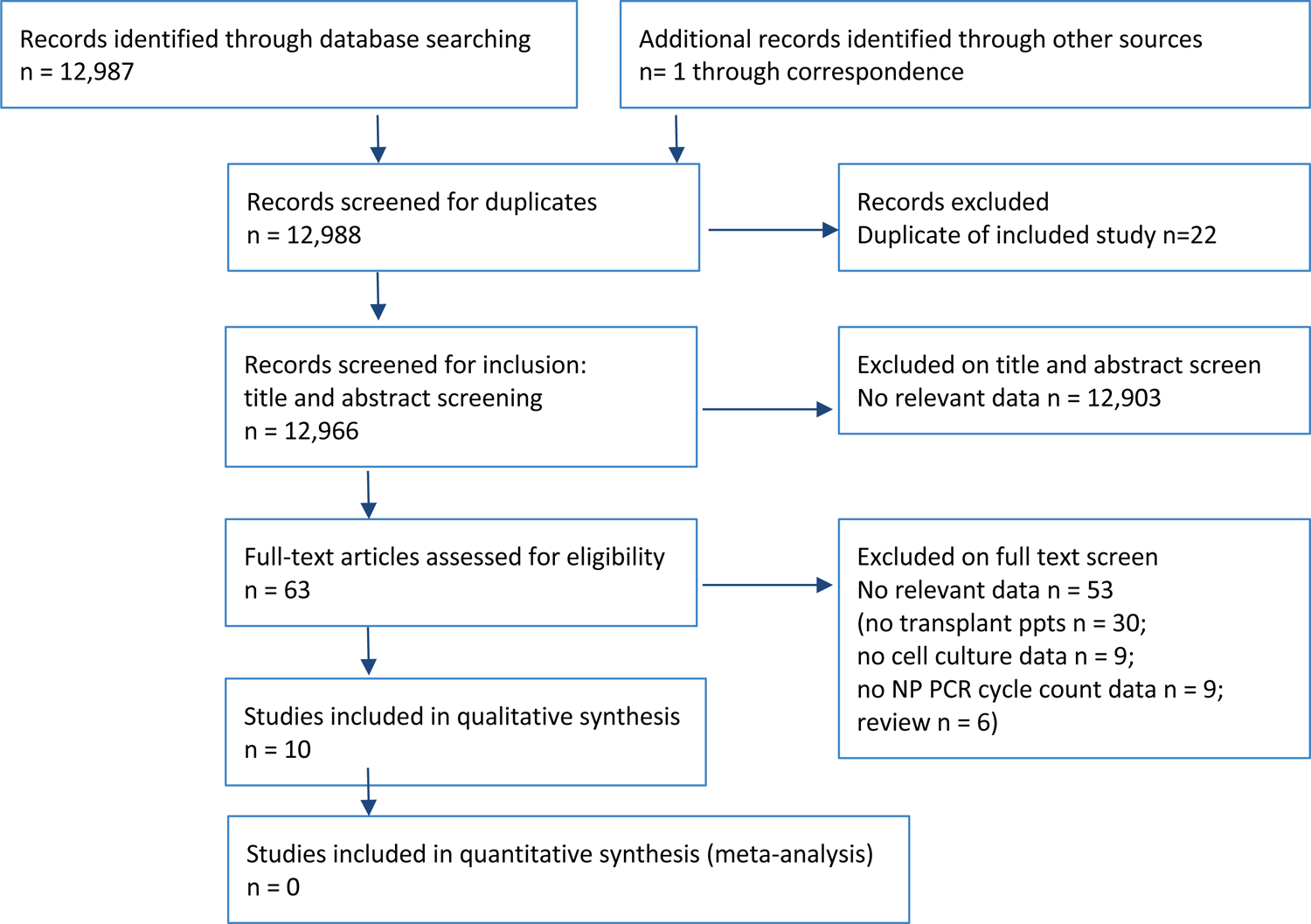
PRISMA flow chart of study screening for inclusion.

A total of 36 were infected with SARS-CoV-2 post-transplant: 21 patients in 3 studies had had kidney transplant ^7 8 12^, 5 patients in 4 studies had had a cardiac transplant^7 10 15 9^, 1 previous bone marrow transplant for multiple myeloma ^14^, 1 liver transplant ^14^ and eight hematopoietic cell transplants^6^. Two patients were infected with SARS-CoV-2 and subsequently underwent transplant: 1 liver transplant ^11^, 1 patient had bilateral lung transplantation after a SARS-CoV-2 infection that severely affected the lungs ^13^.

Typically, patients received ^6^a mixture of antivirals, antibiotics, convalescent plasma and immune suppressants, as reported in Table 1. The clinical course of COVID-19 varied widely amongst the included patients, from mild COVID-19 related symptoms to severe pneumonia and lung failure; no deaths were specifically reported for this group, although deaths were reported for 4 aggregate patients in one study within 30 days of diagnosis^6^. Prescribed treatments reflected the variation in severity.

**Table 1.**
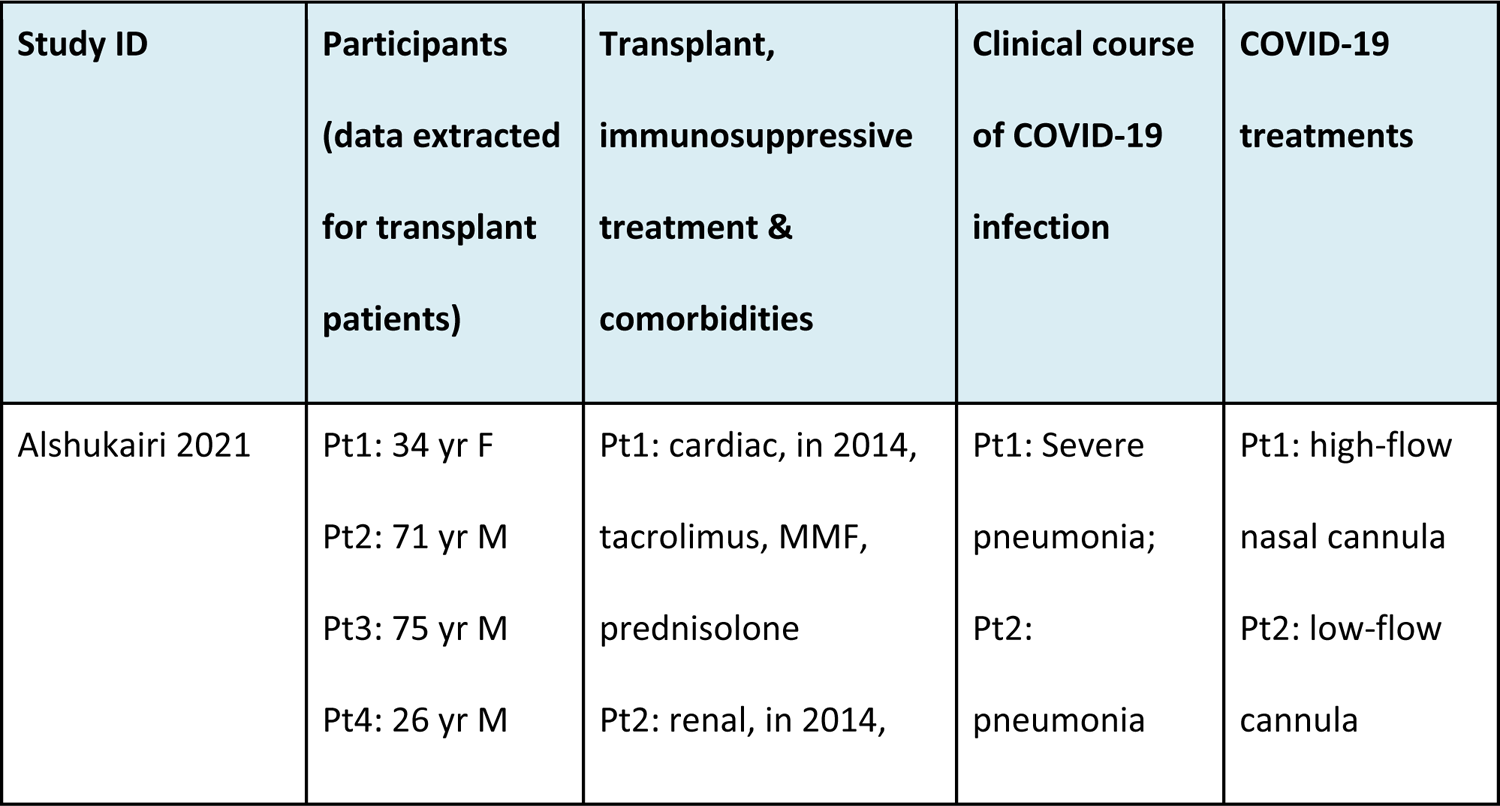
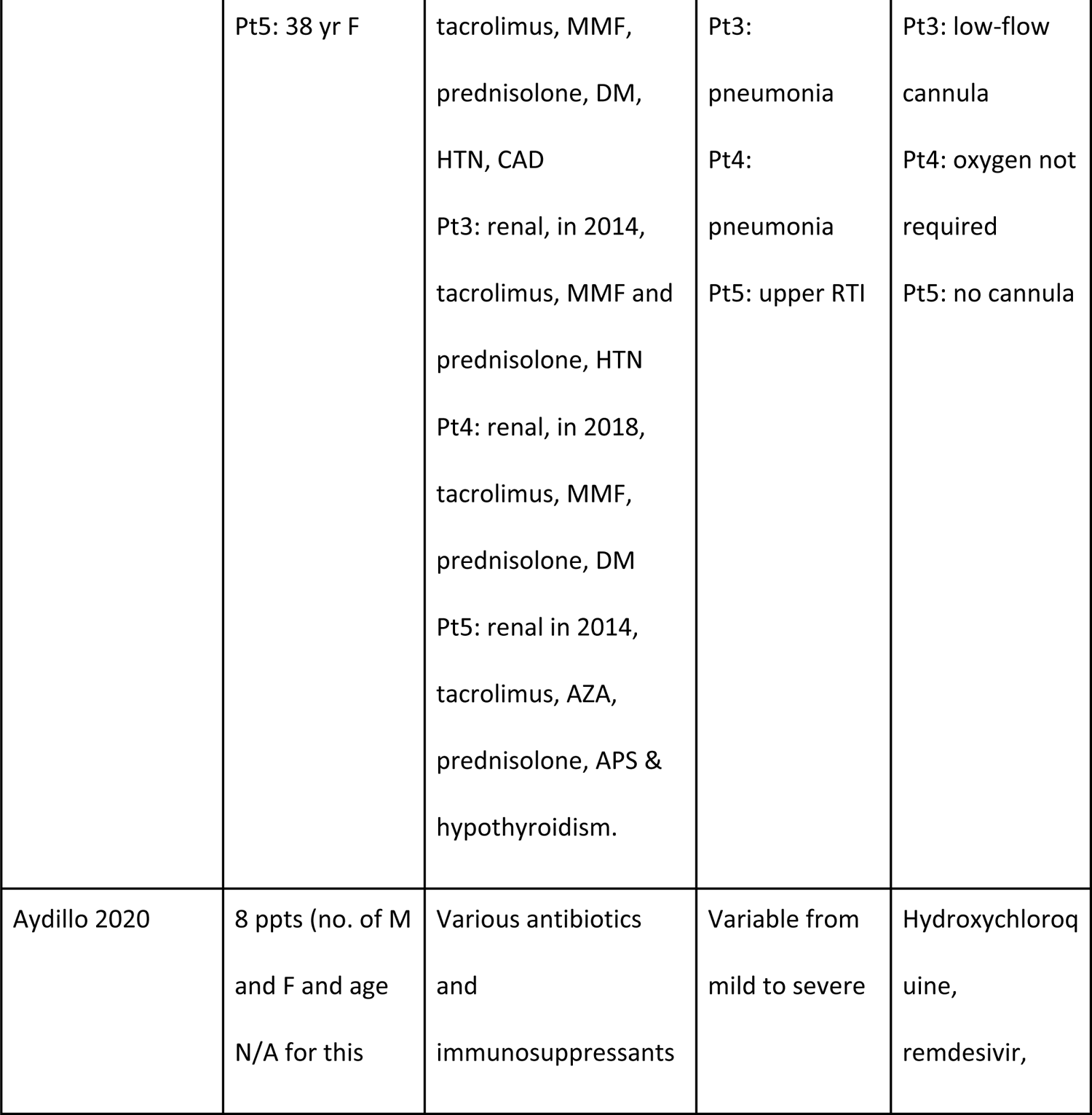
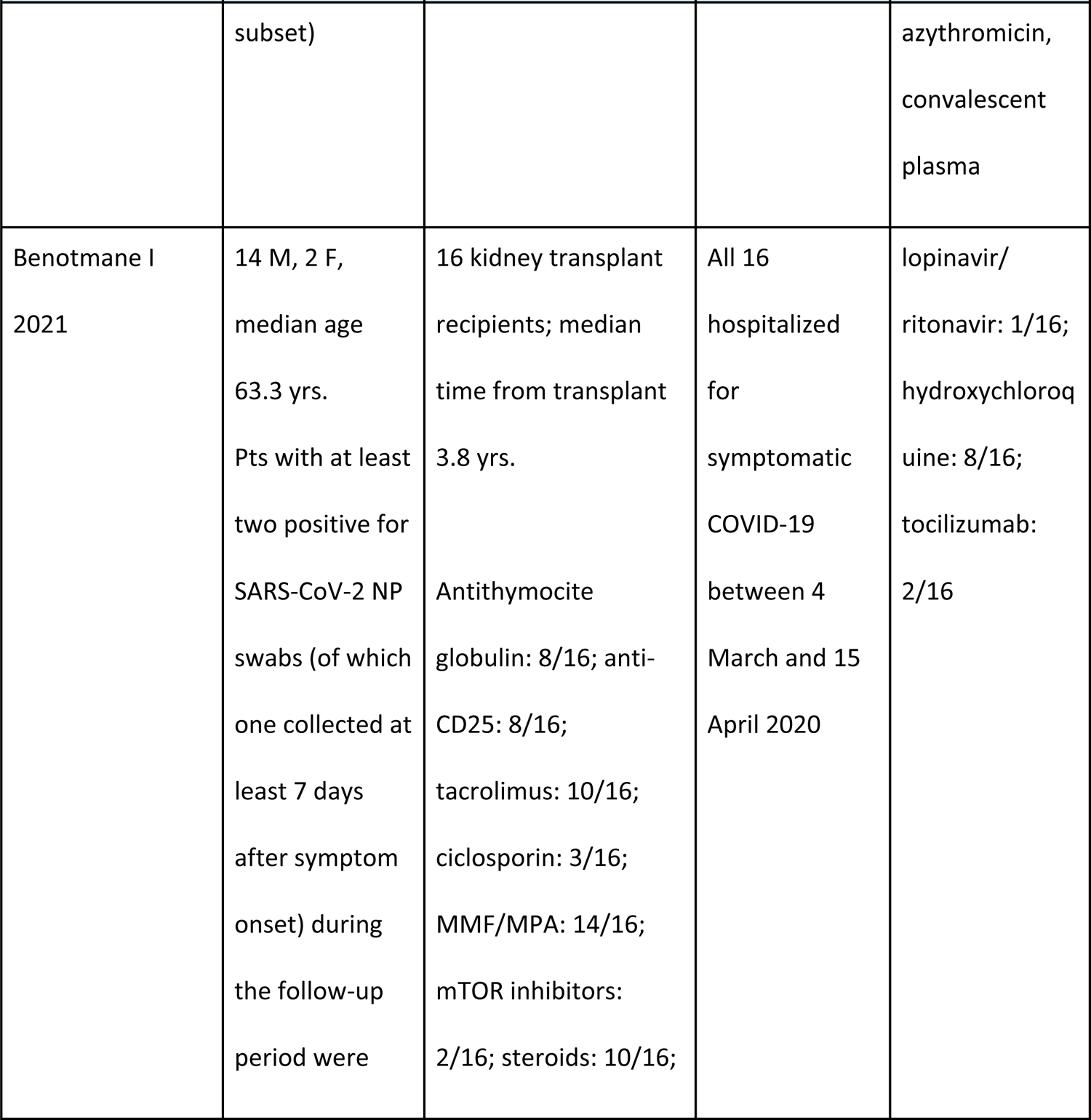
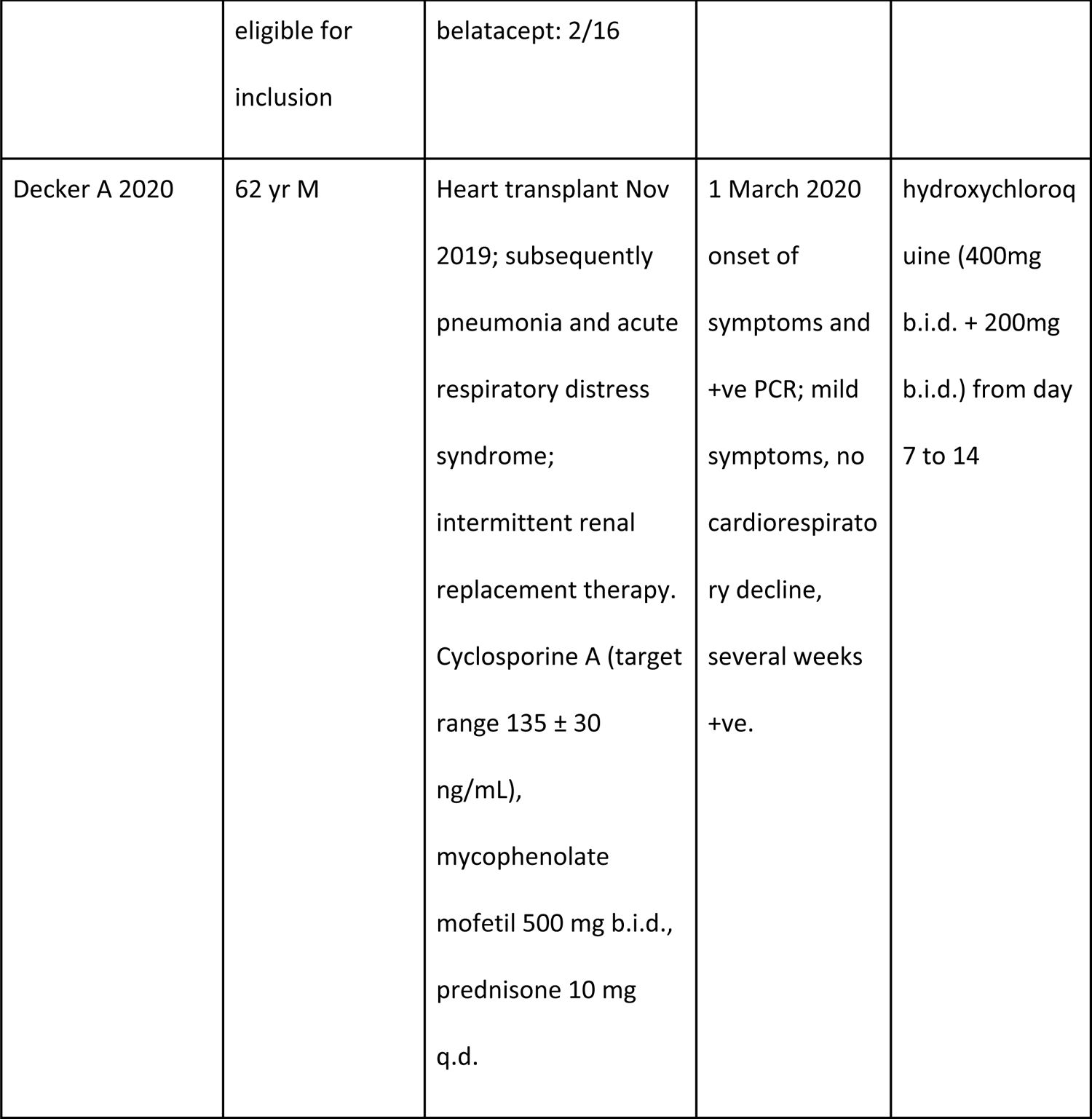
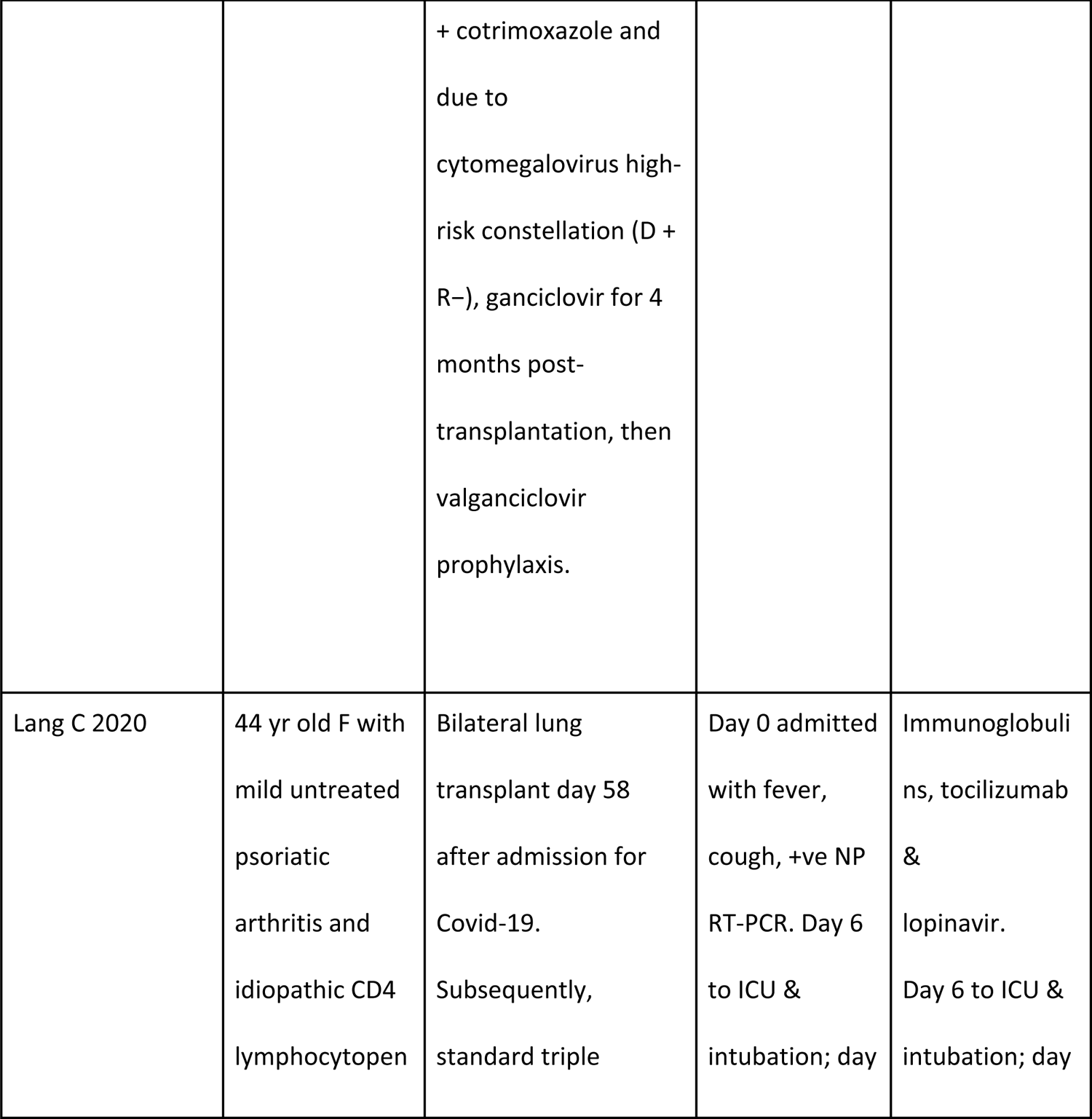
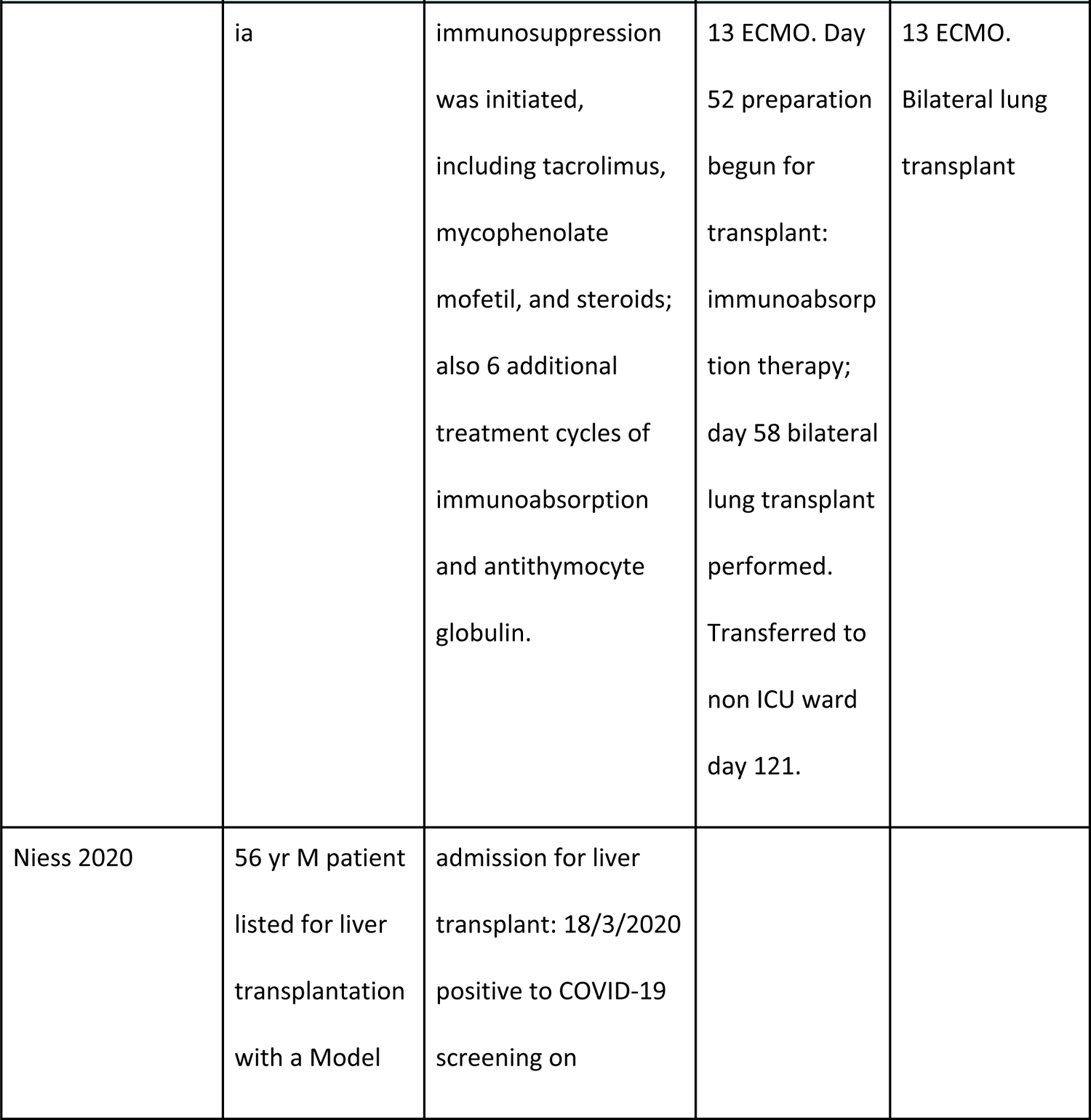
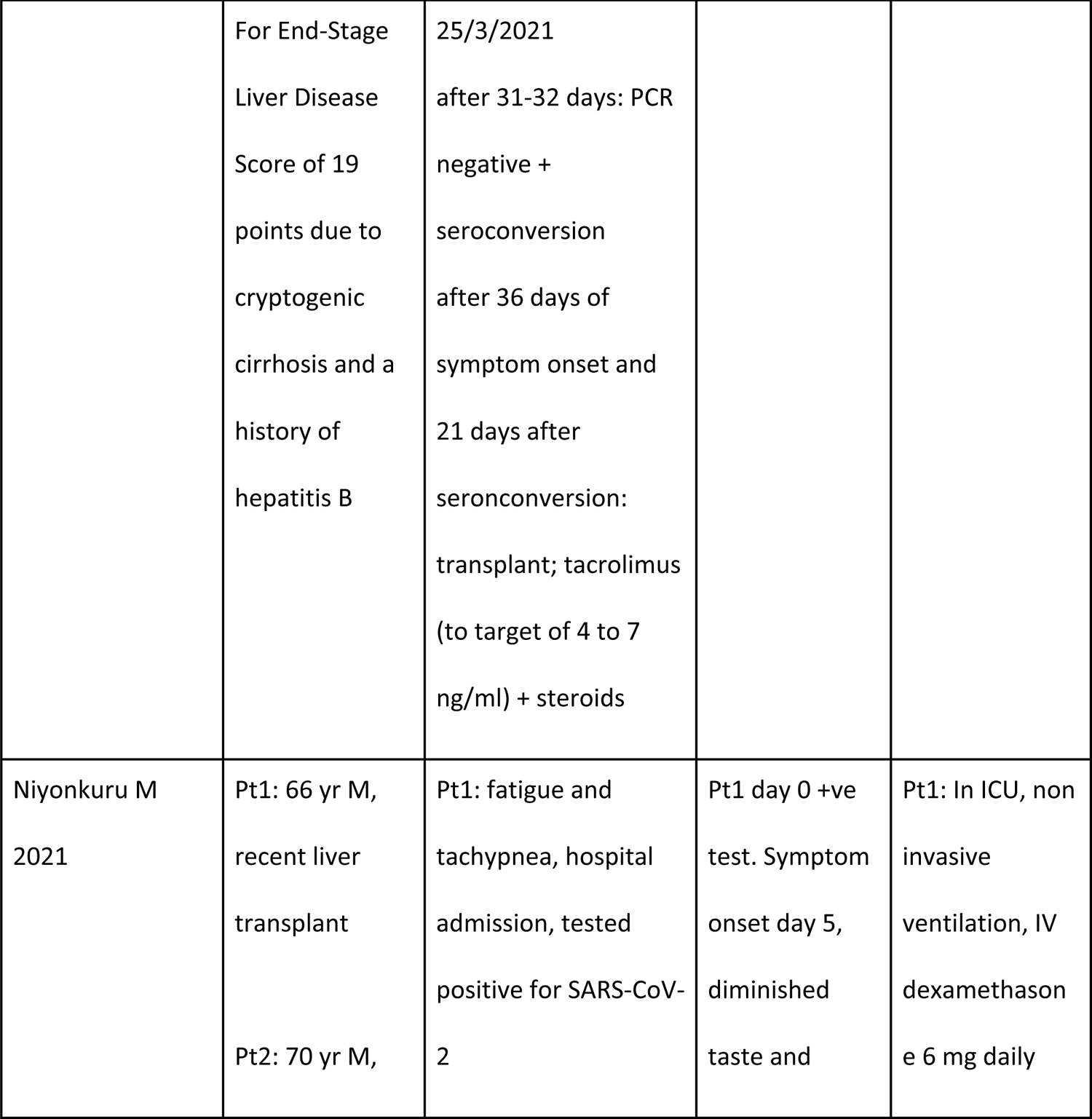
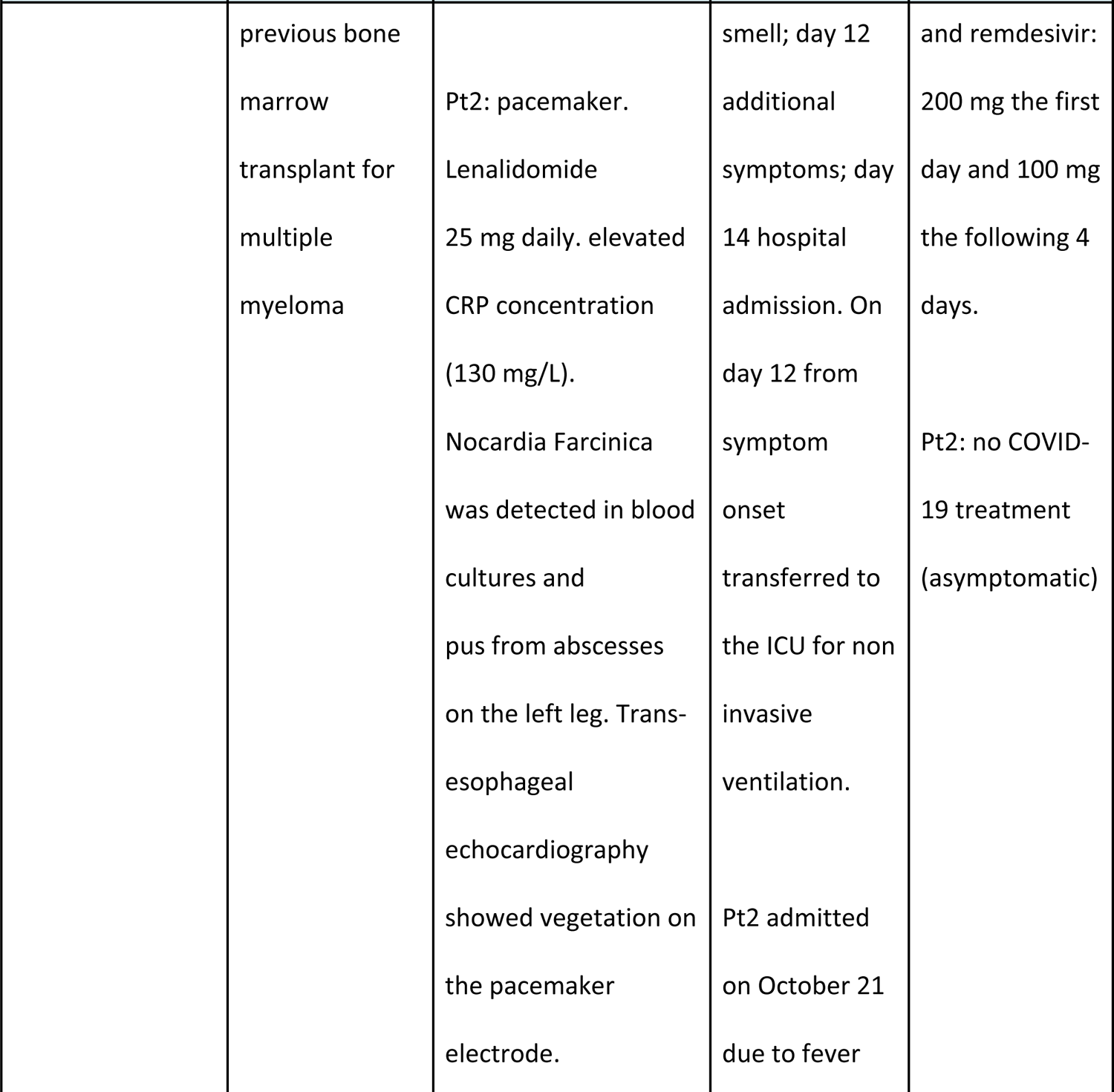
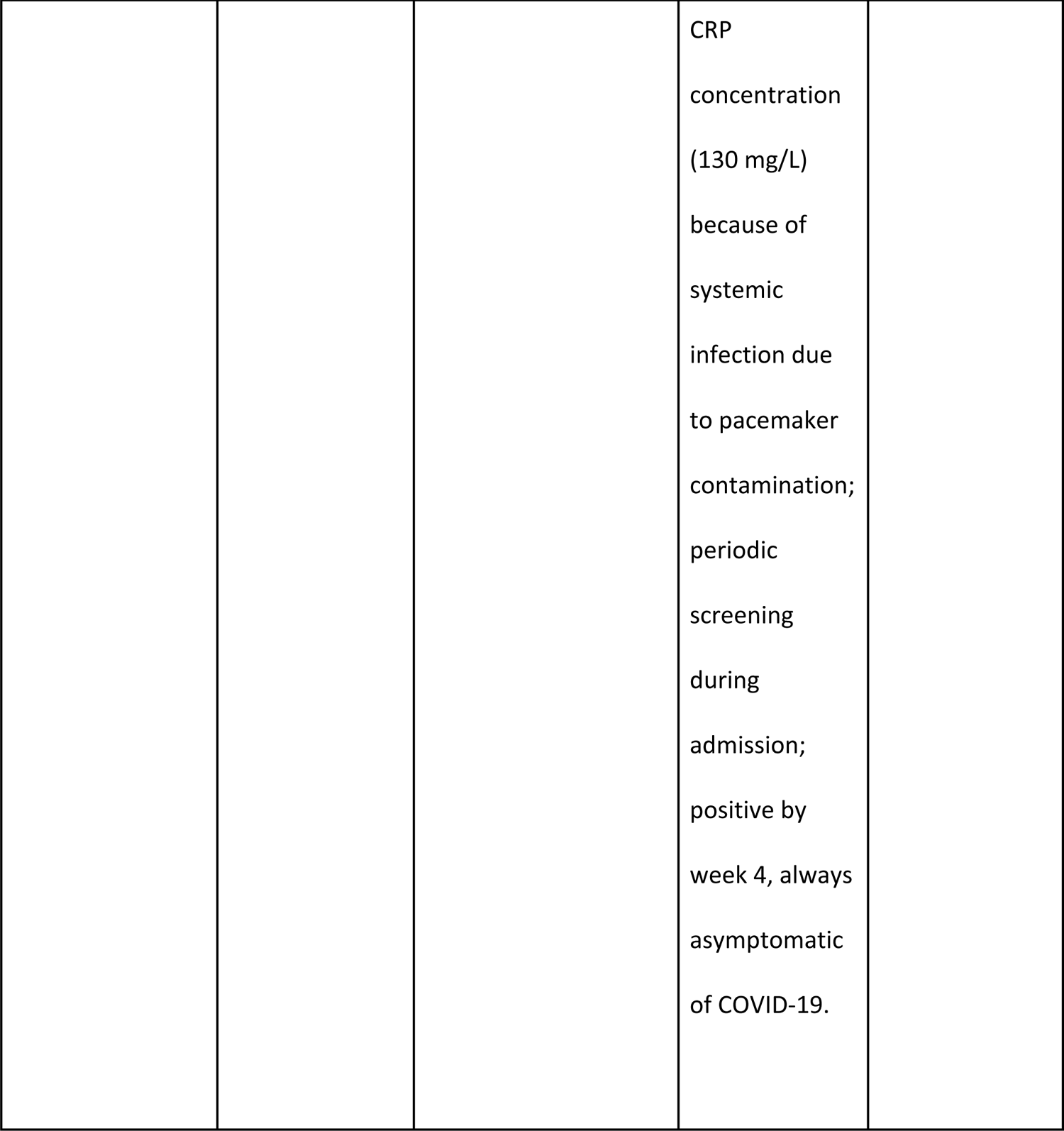
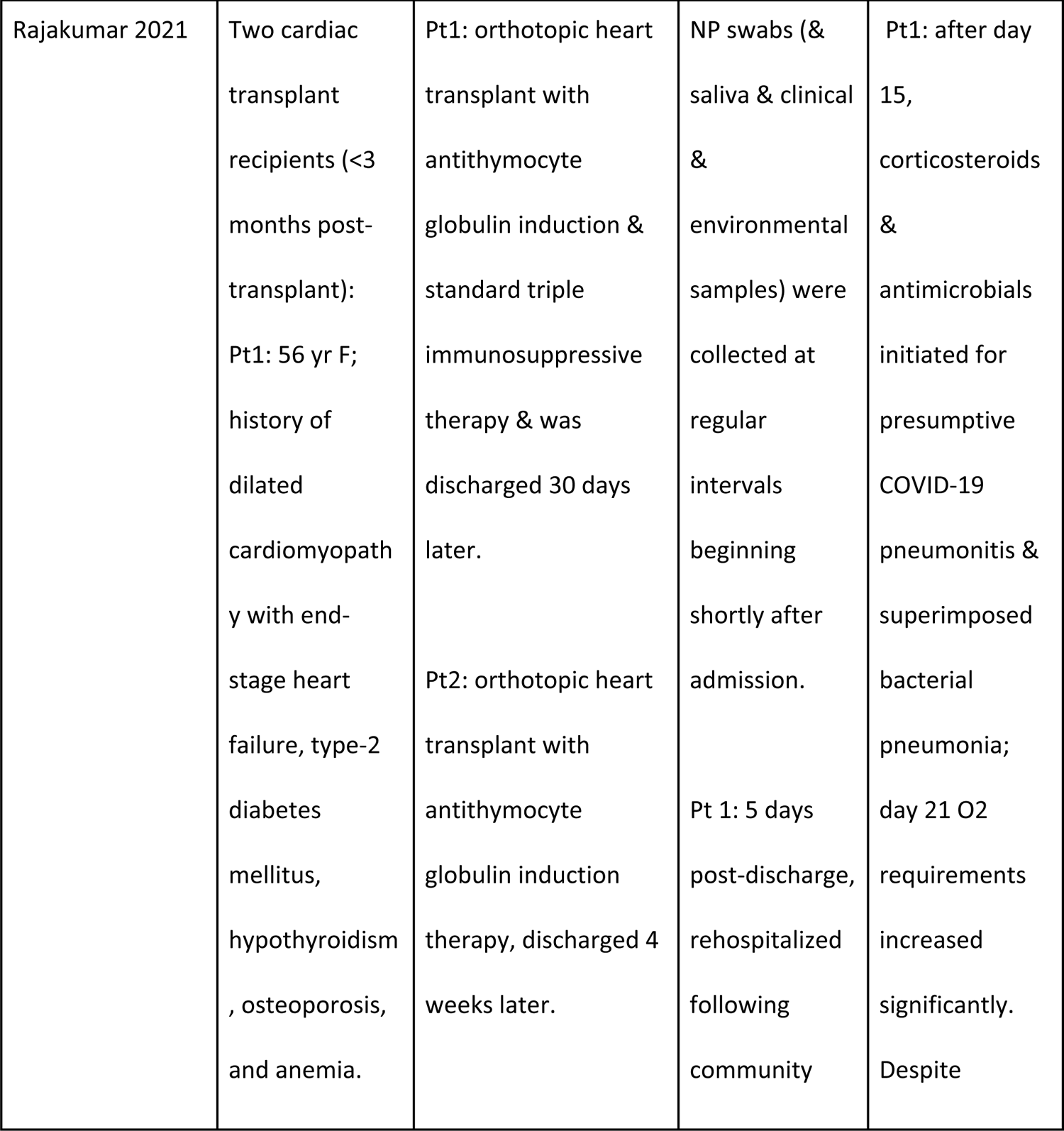
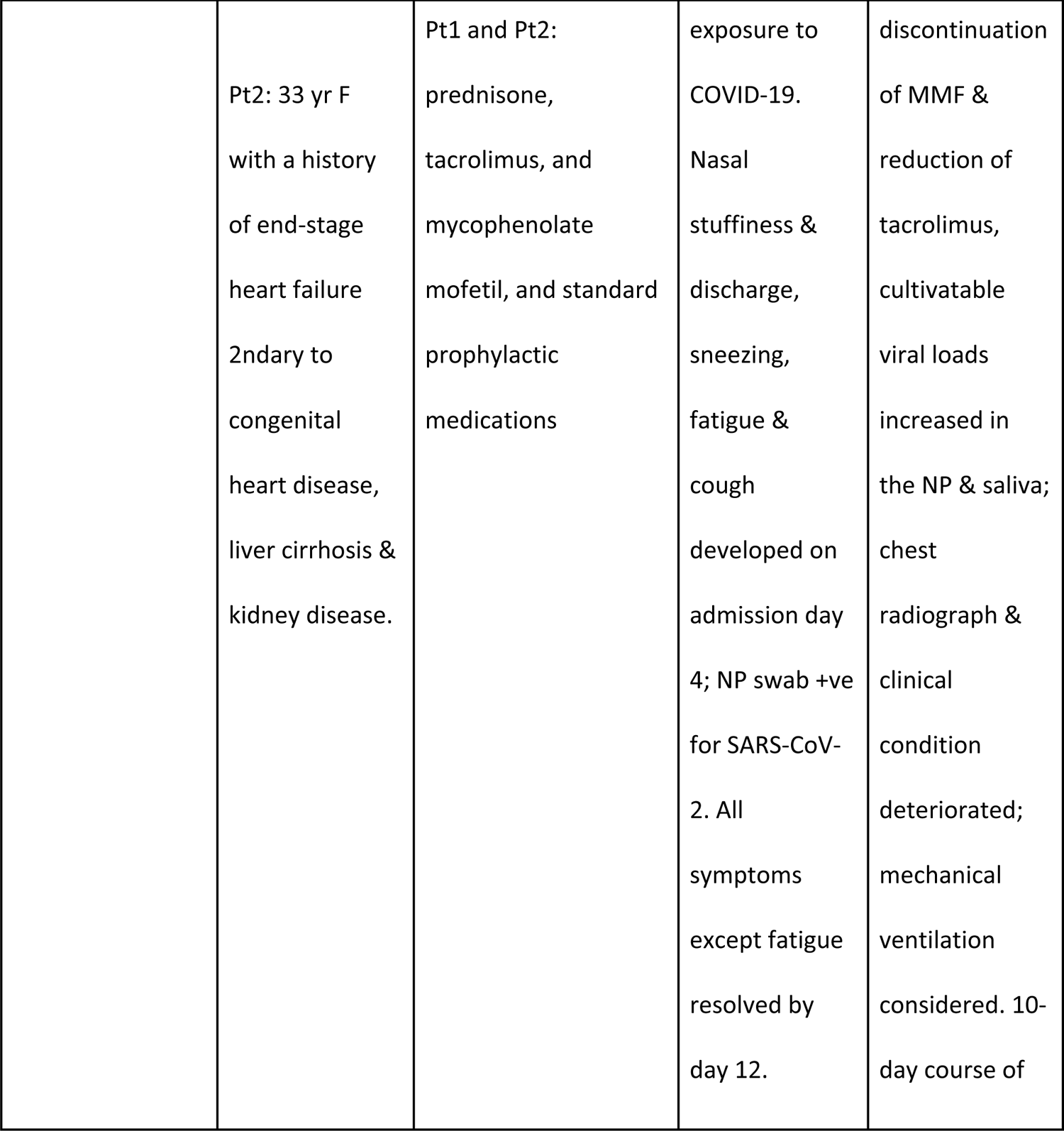
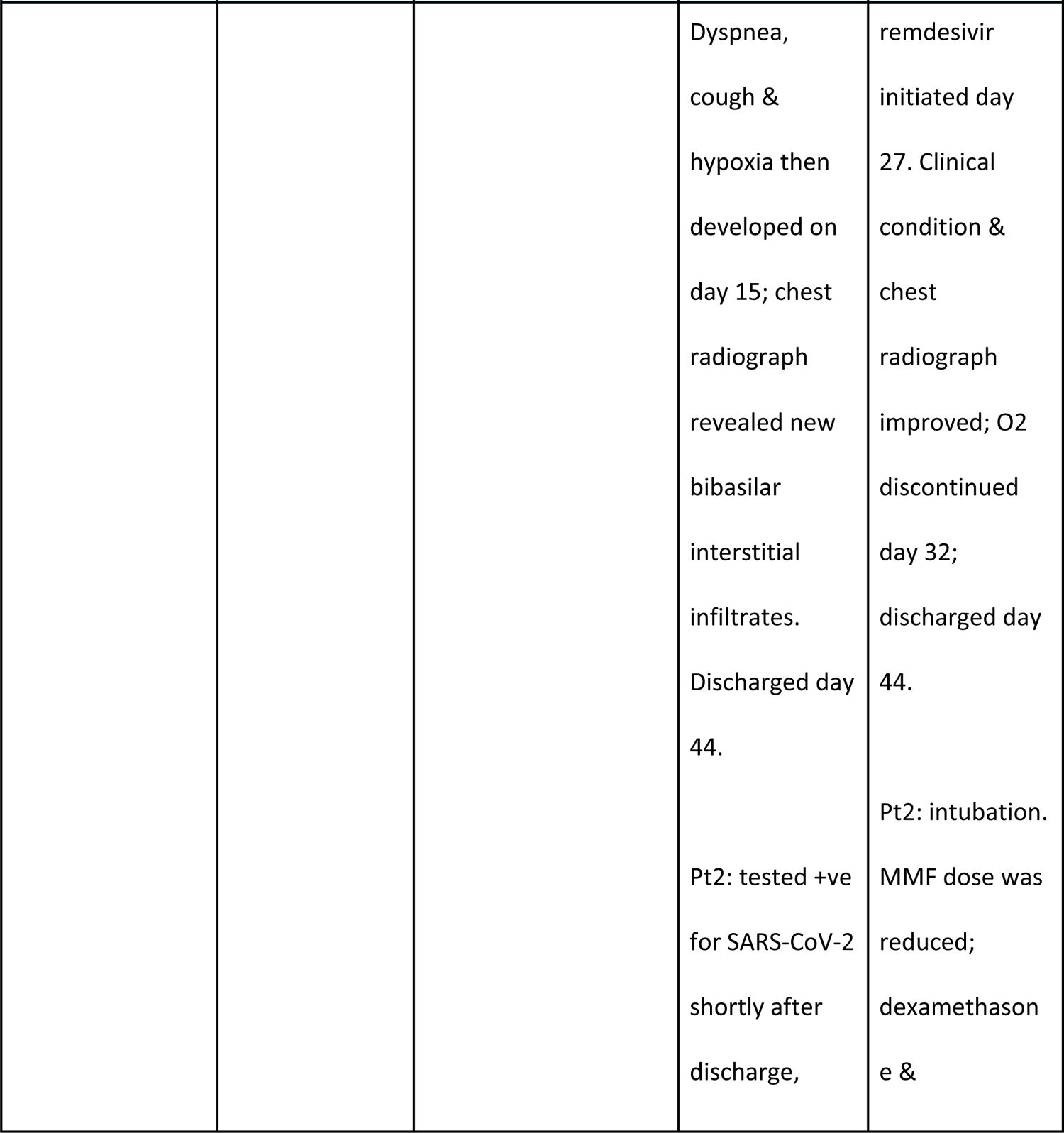
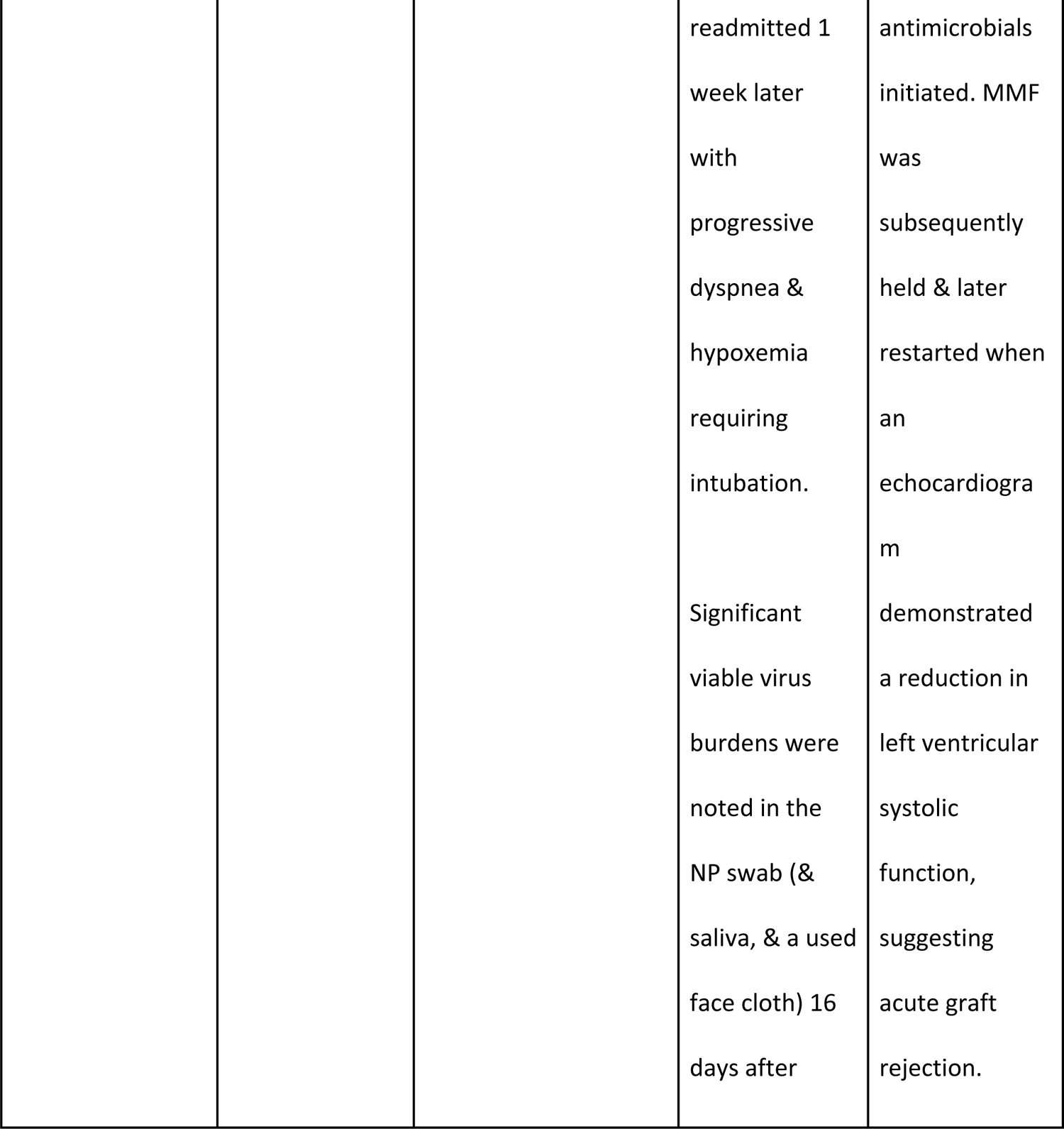
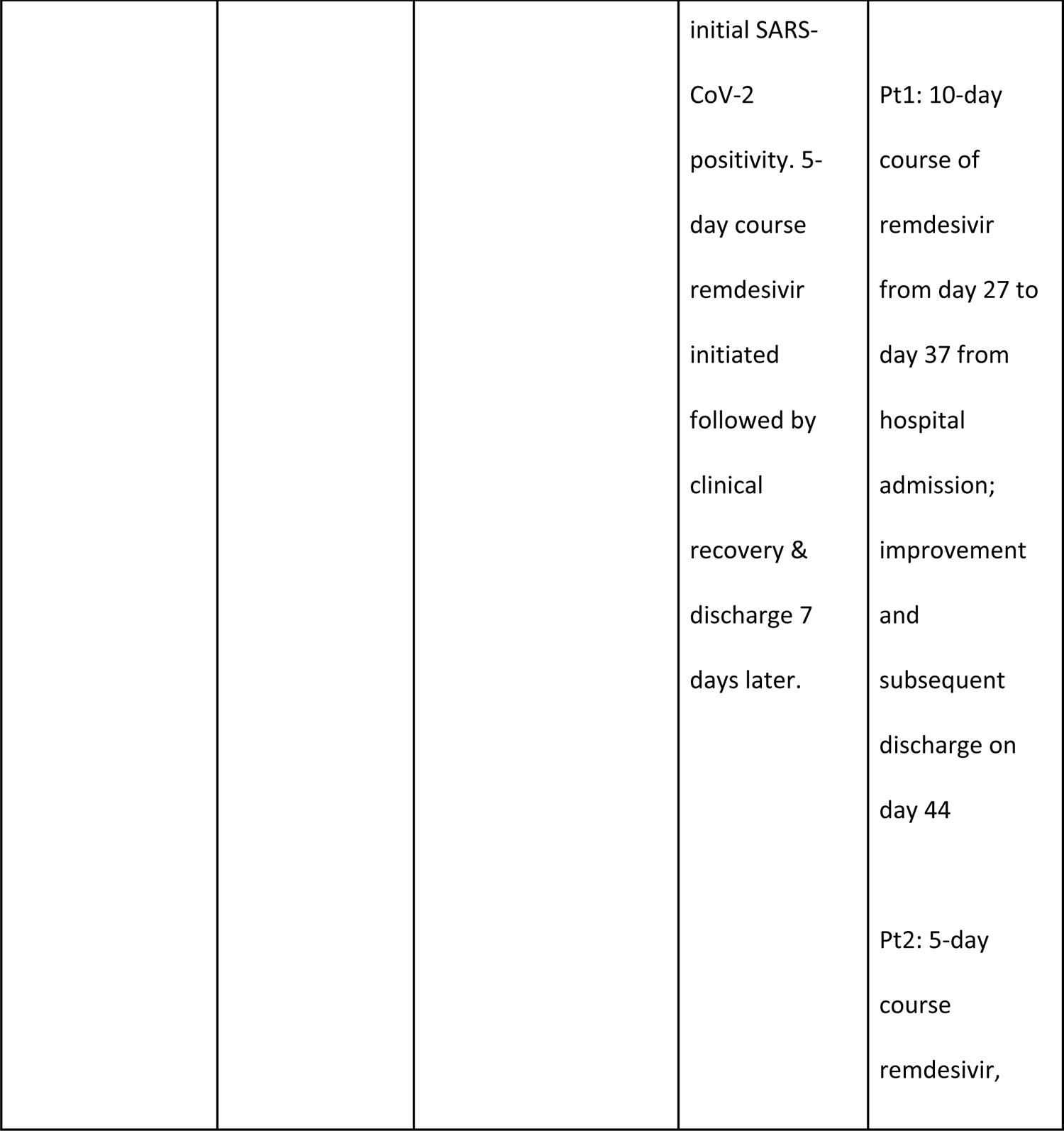
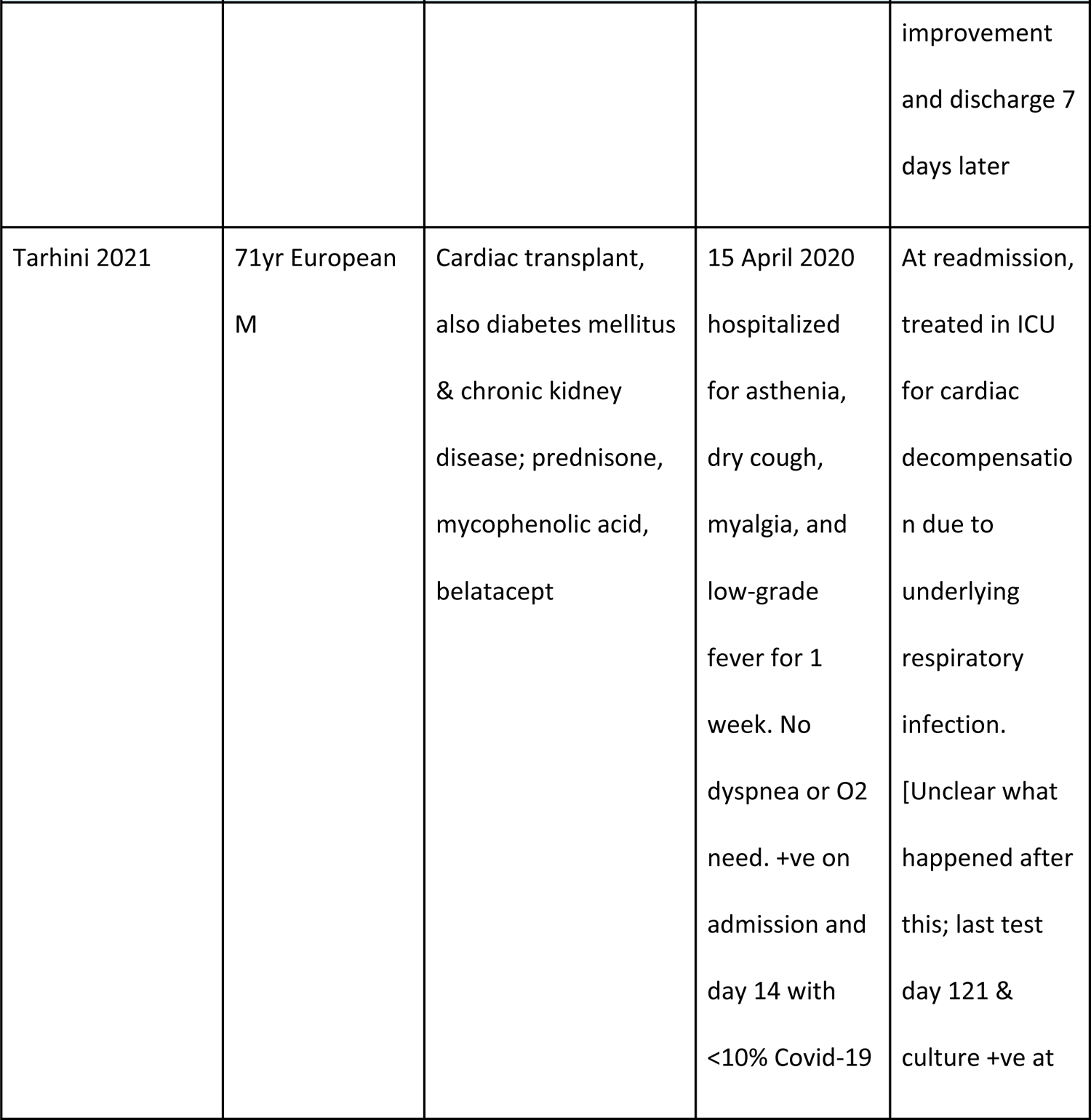
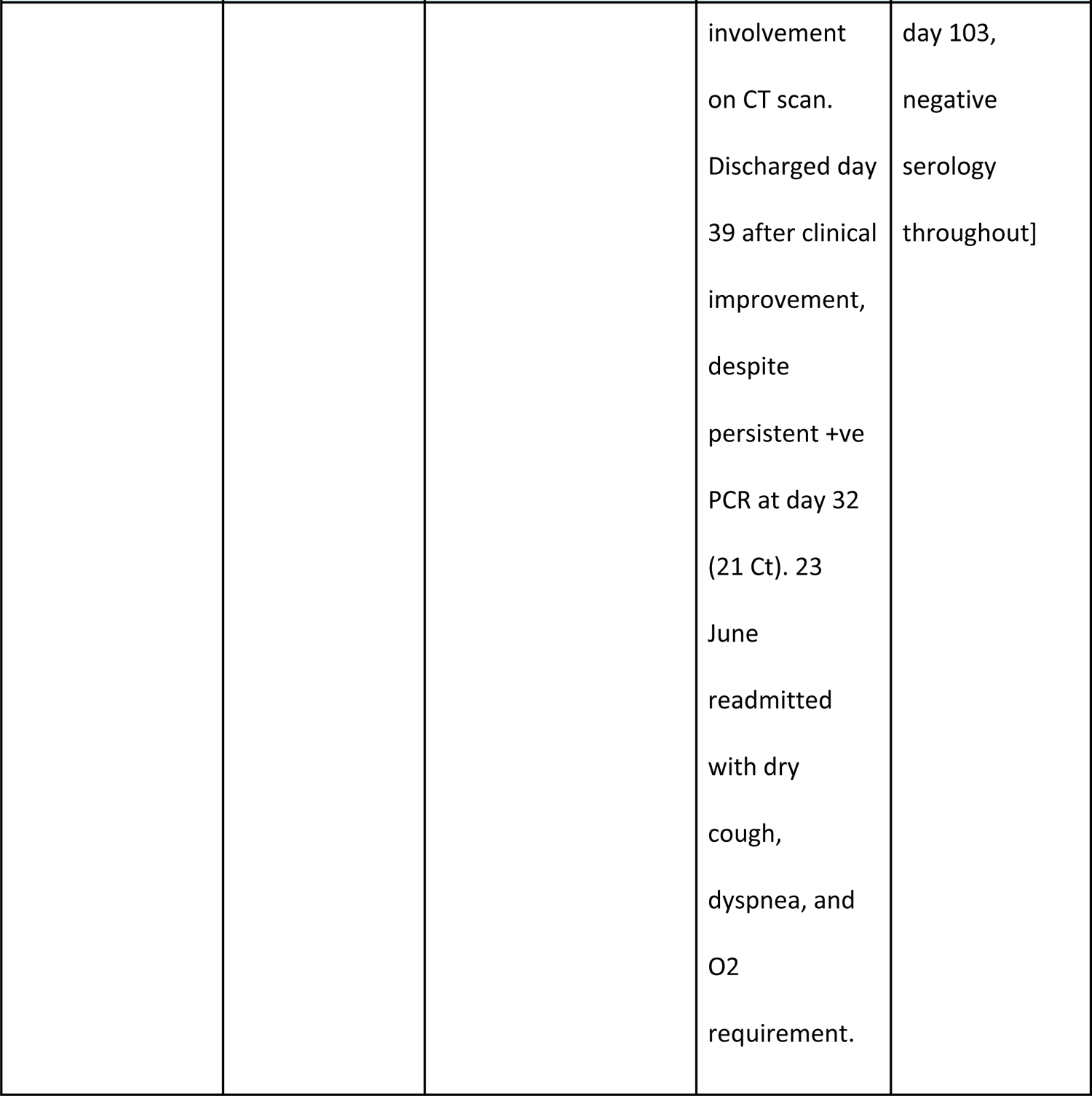
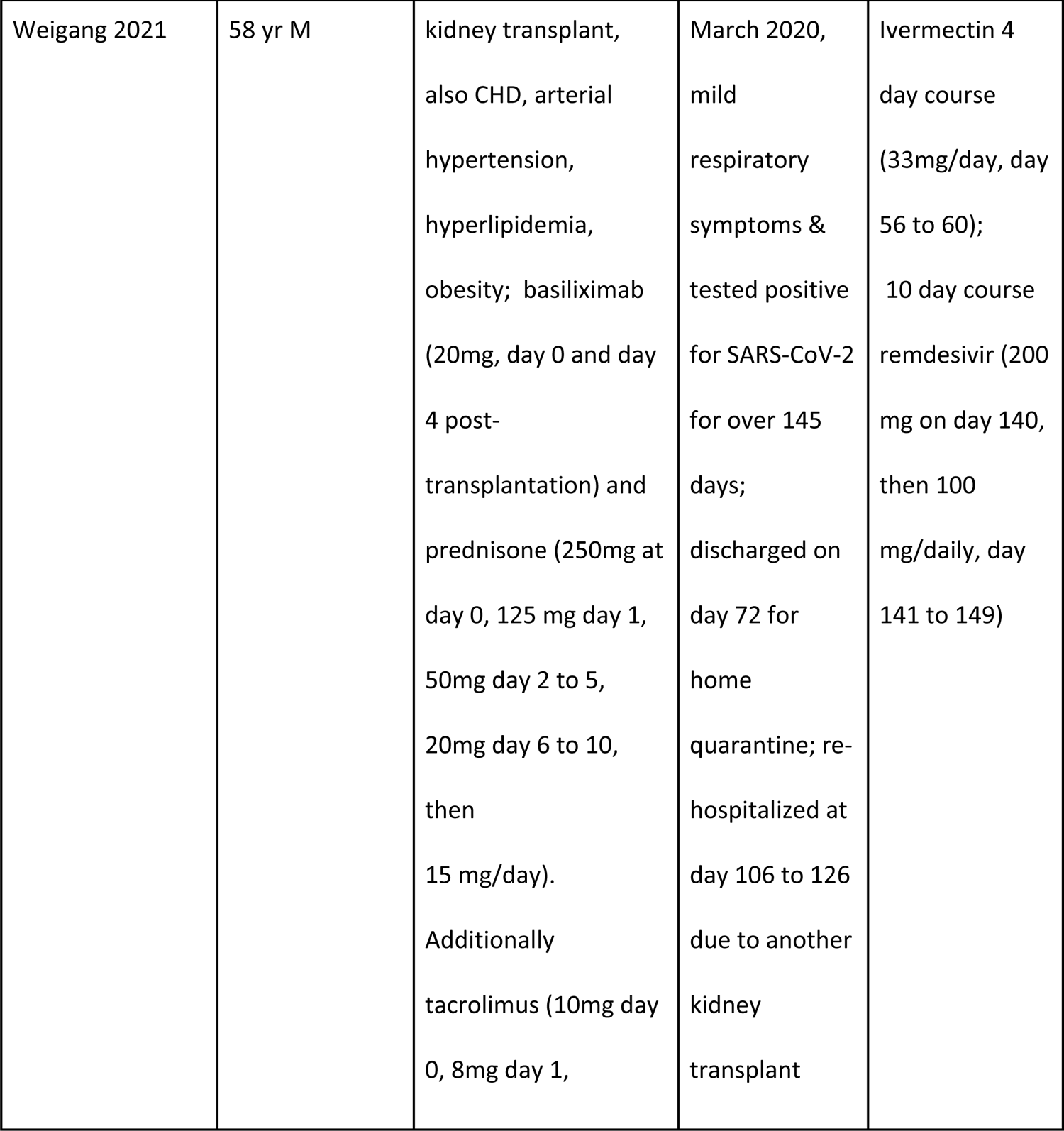
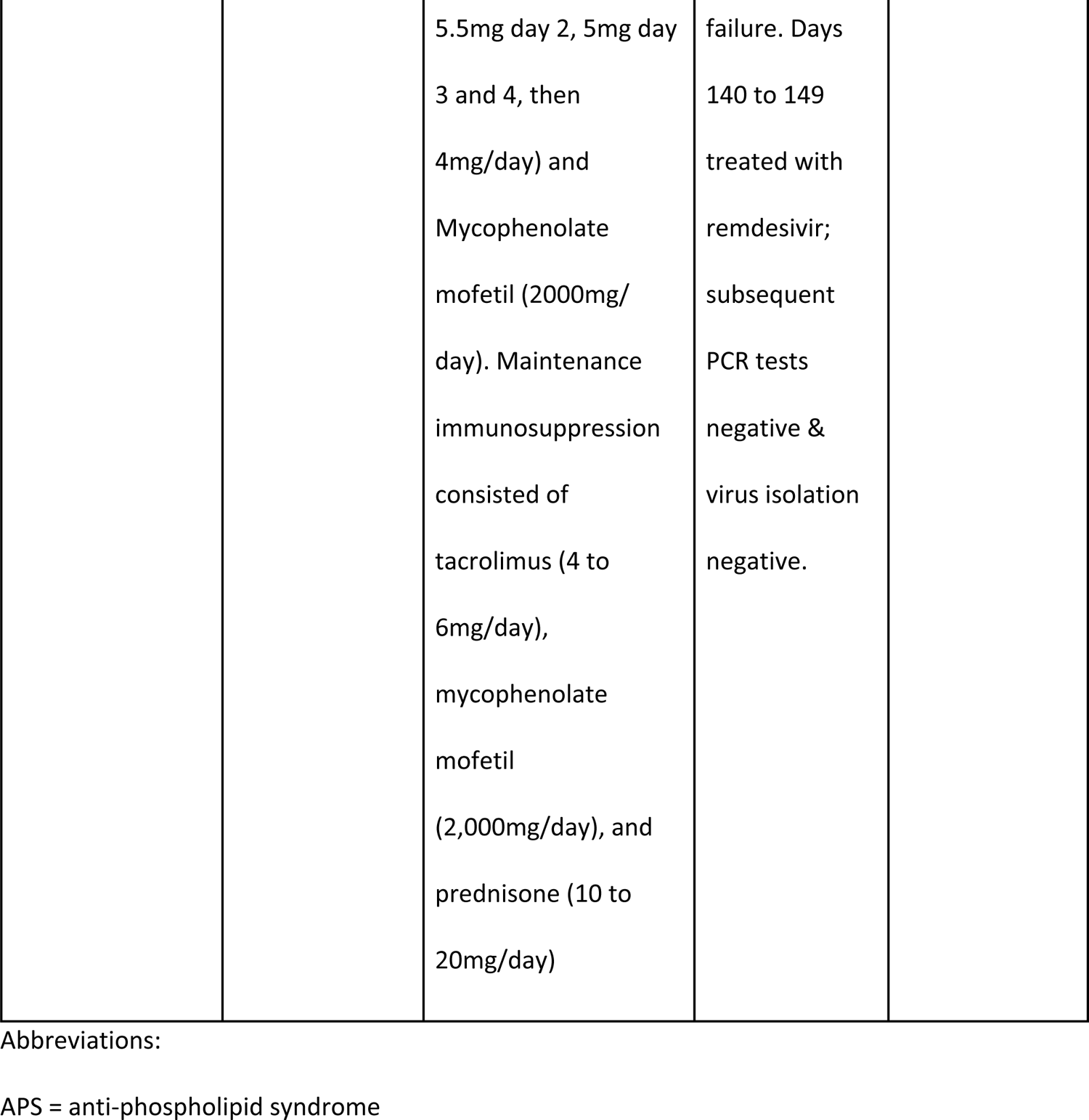
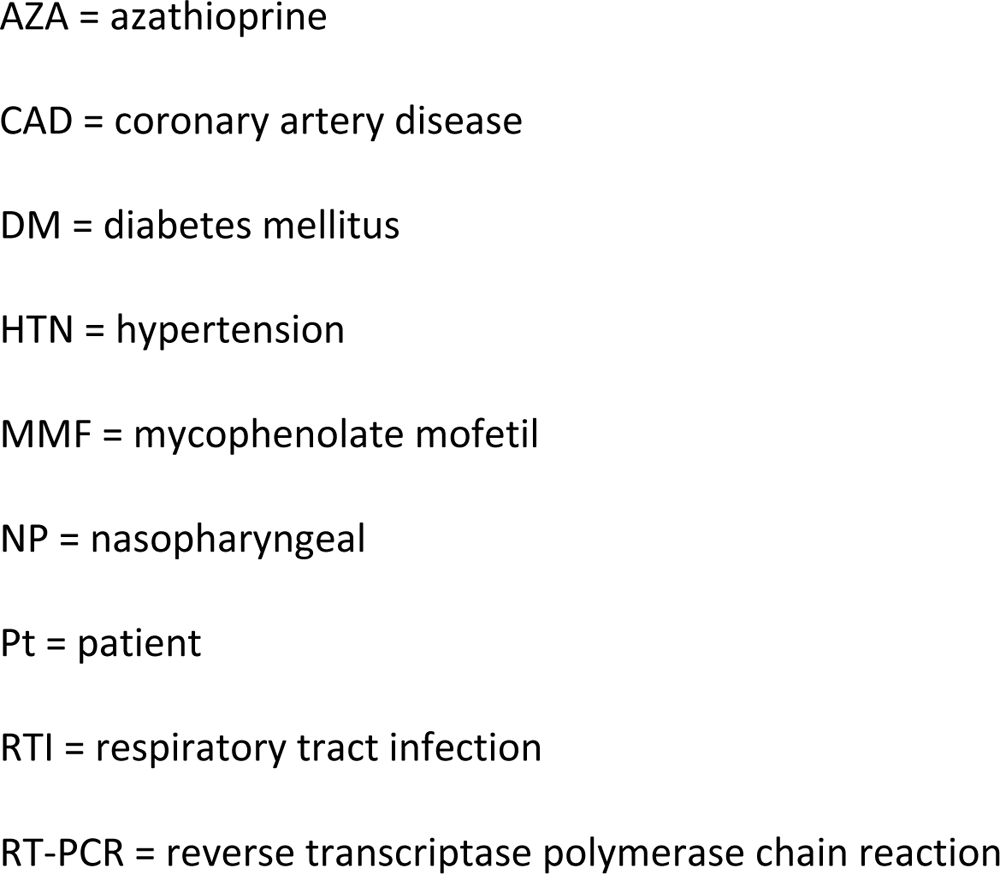
Characteristics of transplant patients in included studies.

| Study ID | Participants<br>(data extracted<br>for transplant<br>patients) | Transplant,<br>immunosuppressive<br>treatment &<br>comorbidities | Clinical course<br>of COVID-19<br>infection | COVID-19<br>treatments |
| --- | --- | --- | --- | --- |
| Alshukairi 2021 | Pt1: 34 yr F<br>Pt2: 71 yr M<br>Pt3: 75 yr M<br>Pt4: 26 yr M | Pt1: cardiac, in 2014,<br>tacrolimus, MMF,<br>prednisolone<br>Pt2: renal, in 2014, | Pt1: Severe<br>pneumonia;<br>Pt2:<br>pneumonia | Pt1: high-flow<br>nasal cannula<br>Pt2: low-flow<br>cannula |
|  | Pt5: 38 yr F | tacrolimus, MMF,<br>prednisolone, DM,<br>HTN, CAD<br>Pt3: renal, in 2014,<br>tacrolimus, MMF and<br>prednisolone, HTN<br>Pt4: renal, in 2018,<br>tacrolimus, MMF,<br>prednisolone, DM<br>Pt5: renal in 2014,<br>tacrolimus, AZA,<br>prednisolone, APS &<br>hypothyroidism. | Pt3:<br>pneumonia<br>Pt4:<br>pneumonia<br>Pt5: upper RTI | Pt3: low-flow<br>cannula<br>Pt4: oxygen not<br>required<br>Pt5: no cannula |
| Aydillo 2020 | 8 ppts (no. of M<br>and F and age<br>N/A for this | Various antibiotics<br>and<br>immunosuppressants | Variable from<br>mild to severe | Hydroxychloroq<br>uine,<br>remdesivir, |
|  | subset) |  |  | azithromycin,<br>convalescent<br>plasma |
| Benotmane I<br>2021 | 14 M, 2 F,<br>median age<br>63.3 yrs.<br>Pts with at least<br>two positive for<br>SARS-CoV-2 NP<br>swabs (of which<br>one collected at<br>least 7 days<br>after symptom<br>onset) during<br>the follow-up<br>period were | 16 kidney transplant<br>recipients; median<br>time from transplant<br>3.8 yrs.<br>Antithymocyte<br>globulin: 8/16; anti-CD25: 8/16;<br>tacrolimus: 10/16;<br>cyclosporin: 3/16;<br>MMF/MPA: 14/16;<br>mTOR inhibitors:<br>2/16; steroids: 10/16; | All 16<br>hospitalized<br>for<br>symptomatic<br>COVID-19<br>between 4<br>March and 15<br>April 2020 | lopinavir/<br>ritonavir: 1/16;<br>hydroxychloroq<br>uine: 8/16;<br>tocilizumab:<br>2/16 |
|  | eligible for<br>inclusion | belatacept: 2/16 |  |  |
| Decker A 2020 | 62 yr M | Heart transplant Nov 2019; subsequently pneumonia and acute respiratory distress syndrome; intermittent renal replacement therapy. Cyclosporine A (target range 135 ± 30 ng/mL), mycophenolate mofetil 500 mg b.i.d., prednisone 10 mg q.d. | 1 March 2020 onset of symptoms and +ve PCR; mild symptoms, no cardiorespiratory decline, several weeks +ve. | hydroxychloroquine (400mg b.i.d. + 200mg b.i.d.) from day 7 to 14 |
|  |  | + cotrimoxazole and<br>due to<br>cytomegalovirus high-<br>risk constellation (D +<br>R-), ganciclovir for 4<br>months post-transplantation, then<br>valganciclovir<br>prophylaxis. |  |  |
| Lang C 2020 | 44 yr old F with<br>mild untreated<br>psoriatic<br>arthritis and<br>idiopathic CD4<br>lymphocytopen | Bilateral lung<br>transplant day 58<br>after admission for<br>Covid-19.<br>Subsequently,<br>standard triple | Day 0 admitted<br>with fever,<br>cough, +ve NP<br>RT-PCR. Day 6<br>to ICU &<br>intubation; day | Immunoglobuli<br>ns, tocilizumab<br>&<br>lopinavir.<br>Day 6 to ICU &<br>intubation; day |
|  | ia | immunosuppression<br>was initiated,<br>including tacrolimus,<br>mycophenolate<br>mofetil, and steroids;<br>also 6 additional<br>treatment cycles of<br>immunoabsorption<br>and antithymocyte<br>globulin. | 13 ECMO. Day<br>52 preparation<br>begun for<br>transplant:<br>immunoabsorp<br>tion therapy;<br>day 58 bilateral<br>lung transplant<br>performed.<br>Transferred to<br>non ICU ward<br>day 121. | 13 ECMO.<br>Bilateral lung<br>transplant |
| Niess 2020 | 56 yr M patient<br>listed for liver<br>transplantation<br>with a Model | admission for liver<br>transplant: 18/3/2020<br>positive to COVID-19<br>screening on |  |  |
|  | For End-Stage<br>Liver Disease<br>Score of 19<br>points due to<br>cryptogenic<br>cirrhosis and a<br>history of<br>hepatitis B | 25/3/2021<br>after 31-32 days: PCR<br>negative +<br>seroconversion<br>after 36 days of<br>symptom onset and<br>21 days after<br>seronconversion:<br>transplant; tacrolimus<br>(to target of 4 to 7<br>ng/ml) + steroids |  |  |
| Niyonkuru M<br>2021 | Pt1: 66 yr M,<br>recent liver<br>transplant<br>Pt2: 70 yr M, | Pt1: fatigue and<br>tachypnea, hospital<br>admission, tested<br>positive for SARS-CoV-2 | Pt1 day 0 +ve<br>test. Symptom<br>onset day 5,<br>diminished<br>taste and | Pt1: In ICU, non<br>invasive<br>ventilation, IV<br>dexamethason<br>e 6 mg daily |
|  | previous bone<br>marrow<br>transplant for<br>multiple<br>myeloma | Pt2: pacemaker.<br>Lenalidomide<br>25 mg daily. elevated<br>CRP concentration<br>(130 mg/L).<br>Nocardia Farcinica<br>was detected in blood<br>cultures and<br>pus from abscesses<br>on the left leg. Trans-<br>esophageal<br>echocardiography<br>showed vegetation on<br>the pacemaker<br>electrode. | smell; day 12<br>additional<br>symptoms; day<br>14 hospital<br>admission. On<br>day 12 from<br>symptom<br>onset<br>transferred to<br>the ICU for non<br>invasive<br>ventilation.<br>Pt2 admitted<br>on October 21<br>due to fever<br>and elevated | and remdesivir:<br>200 mg the first<br>day and 100 mg<br>the following 4<br>days.<br>Pt2: no COVID-19 treatment<br>(asymptomatic) |
|  |  |  | CRP<br>concentration<br>(130 mg/L)<br>because of<br>systemic<br>infection due<br>to pacemaker<br>contamination;<br>periodic<br>screening<br>during<br>admission;<br>positive by<br>week 4, always<br>asymptomatic<br>of COVID-19. |  |
| Rajakumar 2021 | Two cardiac<br>transplant<br>recipients (<3<br>months post-transplant):<br>Pt1: 56 yr F;<br>history of<br>dilated<br>cardiomyopath<br>y with end-stage heart<br>failure, type-2<br>diabetes<br>mellitus,<br>hypothyroidism<br>, osteoporosis,<br>and anemia. | Pt1: orthotopic heart<br>transplant with<br>antithymocyte<br>globulin induction &<br>standard triple<br>immunosuppressive<br>therapy & was<br>discharged 30 days<br>later.<br>Pt2: orthotopic heart<br>transplant with<br>antithymocyte<br>globulin induction<br>therapy, discharged 4<br>weeks later. | NP swabs (&<br>saliva & clinical<br>&<br>environmental<br>samples) were<br>collected at<br>regular<br>intervals<br>beginning<br>shortly after<br>admission.<br>Pt 1: 5 days<br>post-discharge,<br>rehospitalized<br>following<br>community | Pt1: after day<br>15,<br>corticosteroids<br>&<br>antimicrobials<br>initiated for<br>presumptive<br>COVID-19<br>pneumonitis &<br>superimposed<br>bacterial<br>pneumonia;<br>day 21 O2<br>requirements<br>increased<br>significantly.<br>Despite |
|  | Pt2: 33 yr F<br>with a history<br>of end-stage<br>heart failure<br>2ndary to<br>congenital<br>heart disease,<br>liver cirrhosis &<br>kidney disease. | Pt1 and Pt2:<br>prednisone,<br>tacrolimus, and<br>mycophenolate<br>mofetil, and standard<br>prophylactic<br>medications | exposure to<br>COVID-19.<br>Nasal<br>stiffness &<br>discharge,<br>sneezing,<br>fatigue &<br>cough<br>developed on<br>admission day<br>4; NP swab +ve<br>for SARS-CoV-2. All<br>symptoms<br>except fatigue<br>resolved by<br>day 12. | discontinuation<br>of MMF &<br>reduction of<br>tacrolimus,<br>cultivable<br>viral loads<br>increased in<br>the NP & saliva;<br>chest<br>radiograph &<br>clinical<br>condition<br>deteriorated;<br>mechanical<br>ventilation<br>considered. 10-day course of |
|  |  |  | Dyspnea, cough & hypoxia then developed on day 15; chest radiograph revealed new bibasilar interstitial infiltrates. Discharged day 44.<br>Pt2: tested +ve for SARS-CoV-2 shortly after discharge, | remdesivir initiated day 27. Clinical condition & chest radiograph improved; O2 discontinued day 32; discharged day 44.<br>Pt2: intubation. MMF dose was reduced; dexamethasone & |
|  |  |  | readmitted 1 week later with progressive dyspnea & hypoxemia requiring intubation.<br>Significant viable virus burdens were noted in the NP swab (& saliva, & a used face cloth) 16 days after | antimicrobials initiated. MMF was subsequently held & later restarted when an echocardiogram demonstrated a reduction in left ventricular systolic function, suggesting acute graft rejection. |
|  |  |  | initial SARS-CoV-2 positivity. 5-day course remdesivir initiated followed by clinical recovery & discharge 7 days later. | Pt1: 10-day course of remdesivir from day 27 to day 37 from hospital admission; improvement and subsequent discharge on day 44<br>Pt2: 5-day course remdesivir, improvement<br>and discharge 7<br>days later |
| Tarhini 2021 | 71yr European<br>M | Cardiac transplant,<br>also diabetes mellitus<br>& chronic kidney<br>disease; prednisone,<br>mycophenolic acid,<br>belatacept | 15 April 2020<br>hospitalized<br>for asthenia,<br>dry cough,<br>myalgia, and<br>low-grade<br>fever for 1<br>week. No<br>dyspnea or O2<br>need. +ve on<br>admission and<br>day 14 with<br><10% Covid-19 | At readmission,<br>treated in ICU<br>for cardiac<br>decompensatio<br>n due to<br>underlying<br>respiratory<br>infection.<br>[Unclear what<br>happened after<br>this; last test<br>day 121 &<br>culture +ve at |
|  |  |  | involvement<br>on CT scan.<br>Discharged day<br>39 after clinical<br>improvement,<br>despite<br>persistent +ve<br>PCR at day 32<br>(21 Ct). 23<br>June<br>readmitted<br>with dry<br>cough,<br>dyspnea, and<br>O2<br>requirement. | day 103,<br>negative<br>serology<br>throughout] |
| Weigang 2021 | 58 yr M | kidney transplant,<br>also CHD, arterial<br>hypertension,<br>hyperlipidemia,<br>obesity; basiliximab<br>(20mg, day 0 and day<br>4 post-transplantation) and<br>prednisone (250mg at<br>day 0, 125 mg day 1,<br>50mg day 2 to 5,<br>20mg day 6 to 10,<br>then<br>15 mg/day).<br>Additionally<br>tacrolimus (10mg day<br>0, 8mg day 1, | March 2020,<br>mild<br>respiratory<br>symptoms &<br>tested positive<br>for SARS-CoV-2<br>for over 145<br>days;<br>discharged on<br>day 72 for<br>home<br>quarantine; re-<br>hospitalized at<br>day 106 to 126<br>due to another<br>kidney<br>transplant | Ivermectin 4<br>day course<br>(33mg/day, day<br>56 to 60);<br>10 day course<br>remdesivir (200<br>mg on day 140,<br>then 100<br>mg/daily, day<br>141 to 149) |
|  |  | 5.5mg day 2, 5mg day 3 and 4, then 4mg/day) and Mycophenolate mofetil (2000mg/day). Maintenance immunosuppression consisted of tacrolimus (4 to 6mg/day), mycophenolate mofetil (2,000mg/day), and prednisone (10 to 20mg/day) | failure. Days 140 to 149 treated with remdesivir; subsequent PCR tests negative & virus isolation negative. |  |

Abbreviations:
APS = anti-phospholipid syndrome
AZA = azathioprine CAD = coronary artery disease DM = diabetes mellitus HTN = hypertension MMF = mycophenolate mofetil NP = nasopharyngeal Pt = patient RTI = respiratory tract infection RT-PCR = reverse transcriptase polymerase chain reaction

### Quality Assessment

Table 2 reports study quality based on five criteria. Three studies ^15 9 12^ met all five criteria. Follow-up was judged adequate in all studies; in nine studies the reporting of patient characteristics was sufficiently comprehensive^6 8 10 13 11 14 15 9 12^ and clinical information was missing for one study ^7^. Case definition was missing or unclear in four studies^6, 7^ ^10^ ^14^, and methods for RT-PCR testing were unclear for three studies^6 10 13^. The methods used for viral culture were unclear in four studies ^7 10 13 11^ and one study reported using a cell line that has not typically been used to demonstrate SARS-CoV-2 growth - Buffalo green monkey kidney (BGMK) cell line^8^.

**Table 2.** Quality of included studies.

| **Study ID** | **Were the criteria for diagnosing a case clearly reported and appropriate ?** | **Was the reporting of patient/ population characteristic s adequate?** | **Was the study period, including follow-up, sufficient?** | **Were the methods used to obtain RT-PCR results replicable and appropriate?** | **Were the methods used to obtain viral culture results replicable and appropriate?** |
| --- | --- | --- | --- | --- | --- |
| Alshukairi 2021 | Unclear* | No** | Yes | Yes | Unclear |
| Aydillo 2020 | Unclear | Yes | Yes | Unclear | Yes |
| Benotmane I 2021 | Yes | Yes | Yes | Yes | No*** |
| Decker A 2020 | No | Yes | Yes | Unclear | Unclear |
| Lang C 2020 | Yes | Yes | Yes | Unclear | Unclear |
| Niess 2020 | Yes | Yes | Yes | Yes | Unclear |
| Niyonkuru M 2021 | No* | Yes | Yes | Yes | Yes |
| Rajakumar 2021 | Yes | Yes | Yes | Yes | Yes |
| Tarhini 2021 | Yes | Yes | Yes | Yes | Yes |
| Weigang 2021 | Yes | Yes | Yes | Yes | Yes |

\*case definition unclear, article reports positive RT-PCR, but Ct cut-off not reported.
\*\* data on clinical symptoms lacking
\*\*\*The cell line used was not one that is demonstrated to support SARS-CoV-2 growth.
Therefore the cell culture results are not reliable.

### Results of the studies

The results are reported in Table 1 and Table 3. The clinical course of infection was highly variable (Tables and 1 and 3, and Figures 2 and 3). The time from transplant to infection varied from days to years (see for example Aydillo et al^6^ and Rajakumar et al^15^). Sampling schedules varied between studies, with no regular timetable of testing taking place, so results for PCR and viral culture are available for different time points in a patient’s clinical course and with different gaps in time between samples being taken.

**Figure 2.**
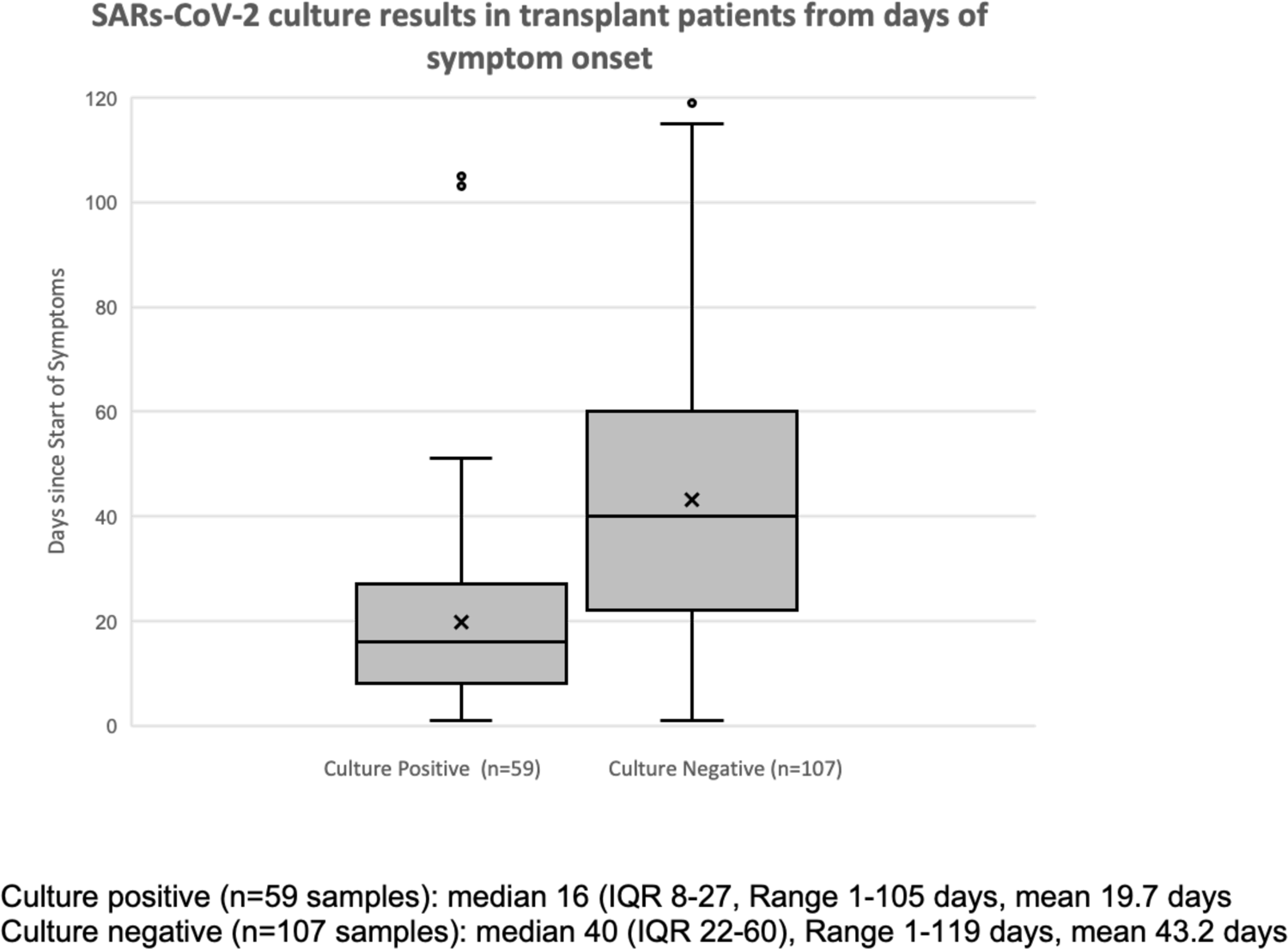
SARs-CoV-2 culture results in transplant patients from days of symptom onset.

**Figure 3a.**
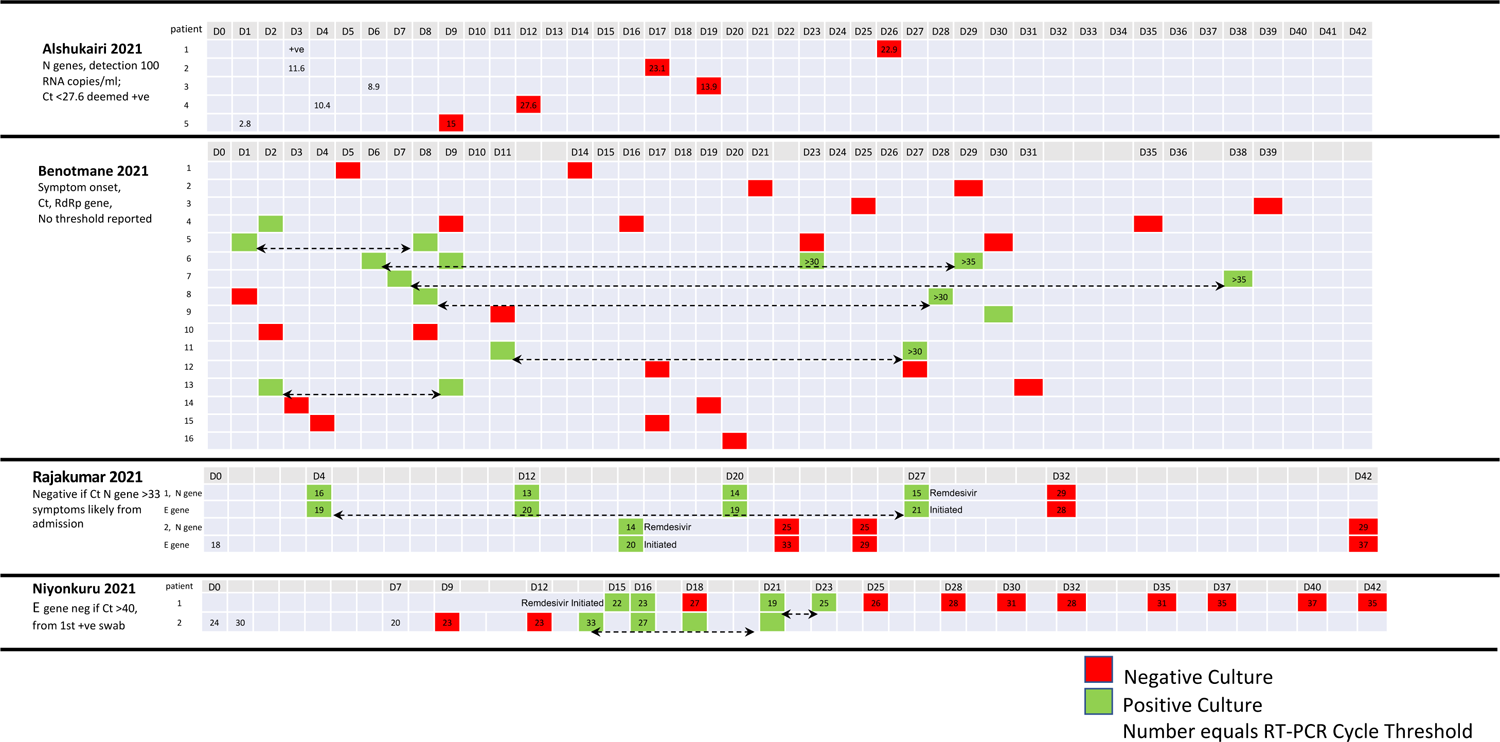
**Figure 3.** Duration of infectivity as indicated by viral culture and corresponding PCR cycle counts/log copies among transplant recipients. Timings of positive culture results in Transplant Patients by duration of symptoms and Ct results Alshukairi ^[7]^, Benotmane ^[8]^, Rajakumar ^[15]^ & Niyonkuru ^[14]^ (days 1-42)

**Figure 3b.**
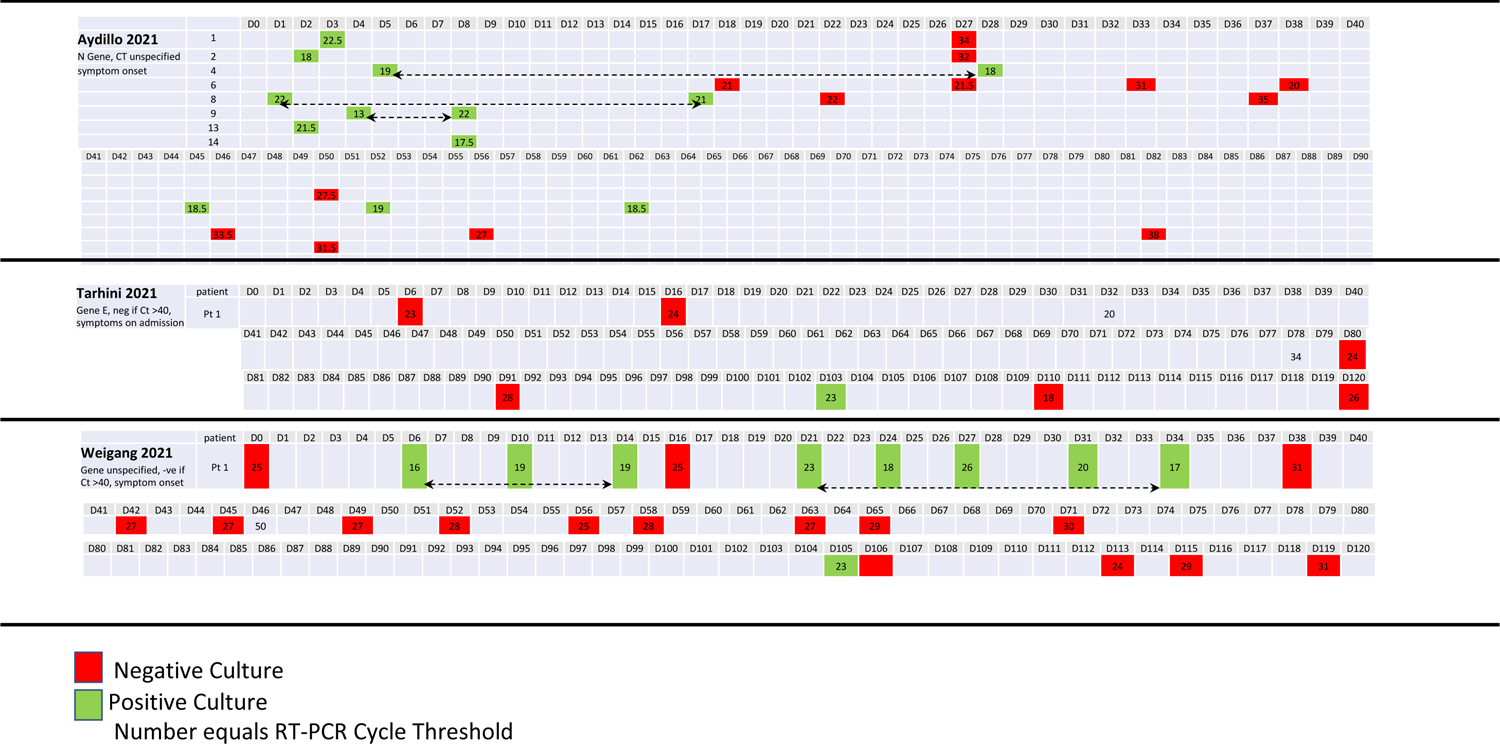
Timings of positive culture results in Transplant Patients by duration of symptoms and Ct results Aydillo ^[6]^ (day 1-90), Tarhini ^[9]^ & Weigang ^[12]^ (day 1-120)

**Figure 3c.**
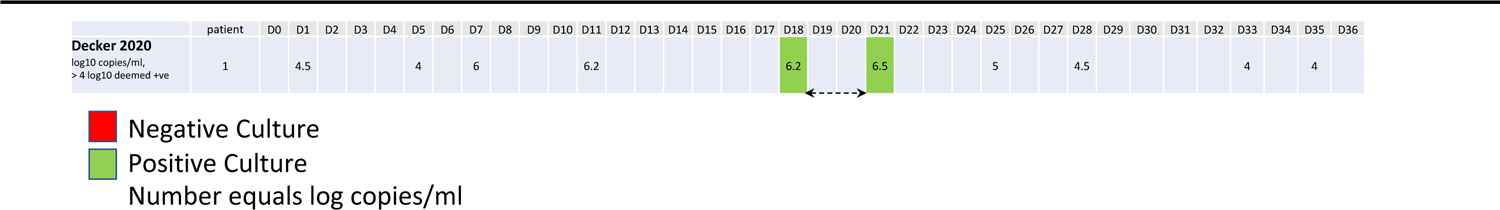
Timings of positive culture results in Transplant Patients by duration of symptoms and log10 copies/ml results Decker ^[10]^ (days 1-36)

**Table 3.**
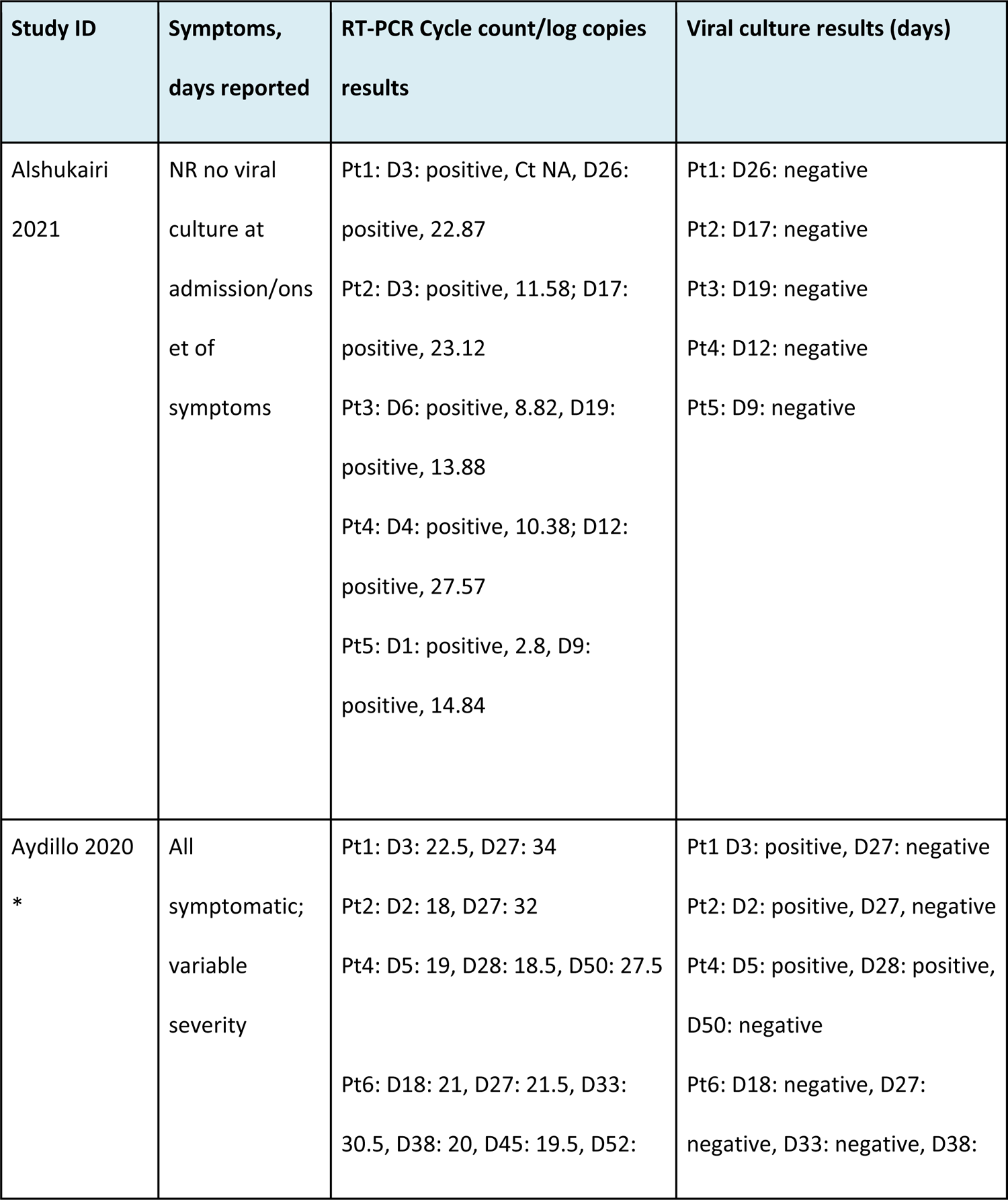
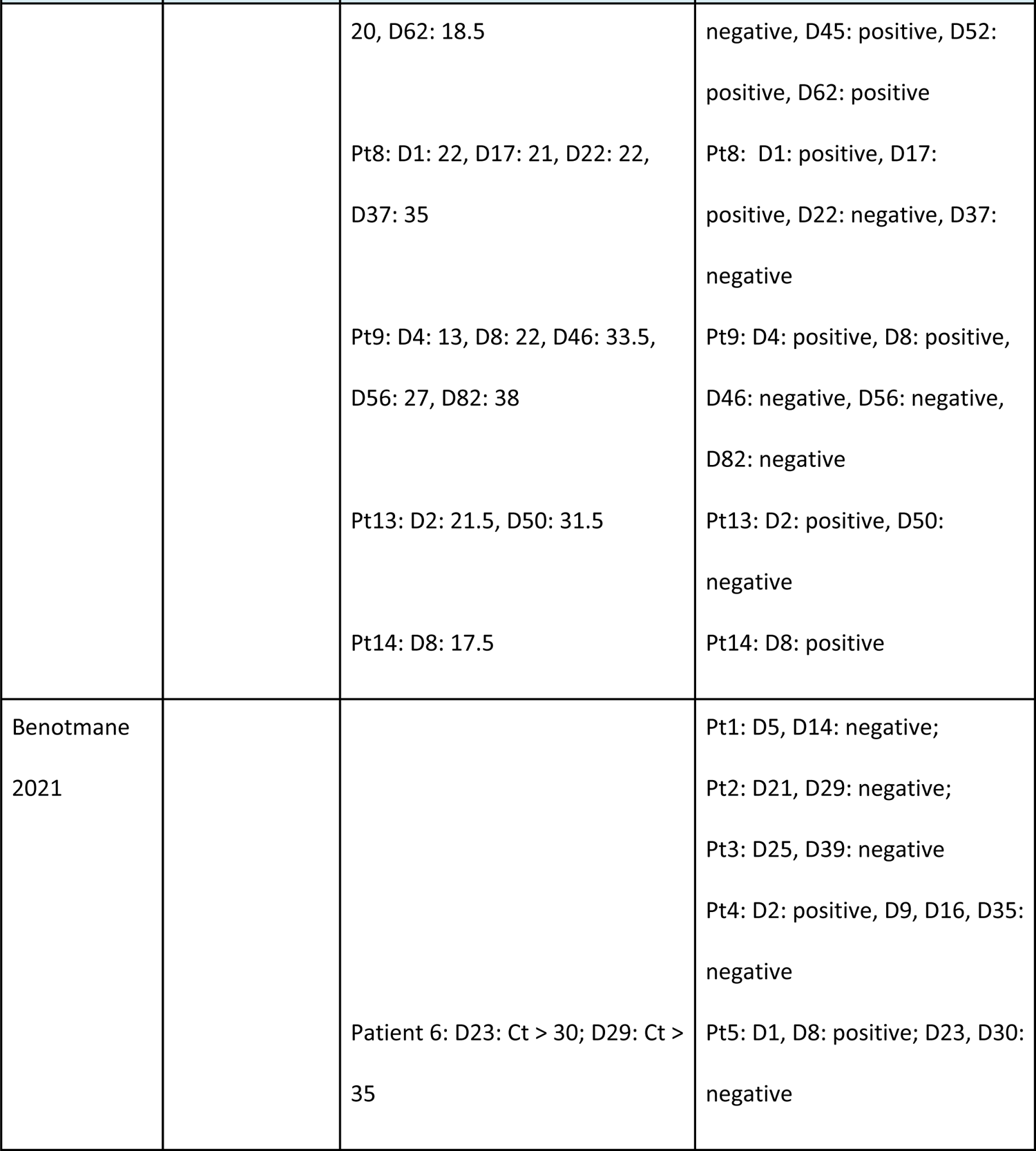
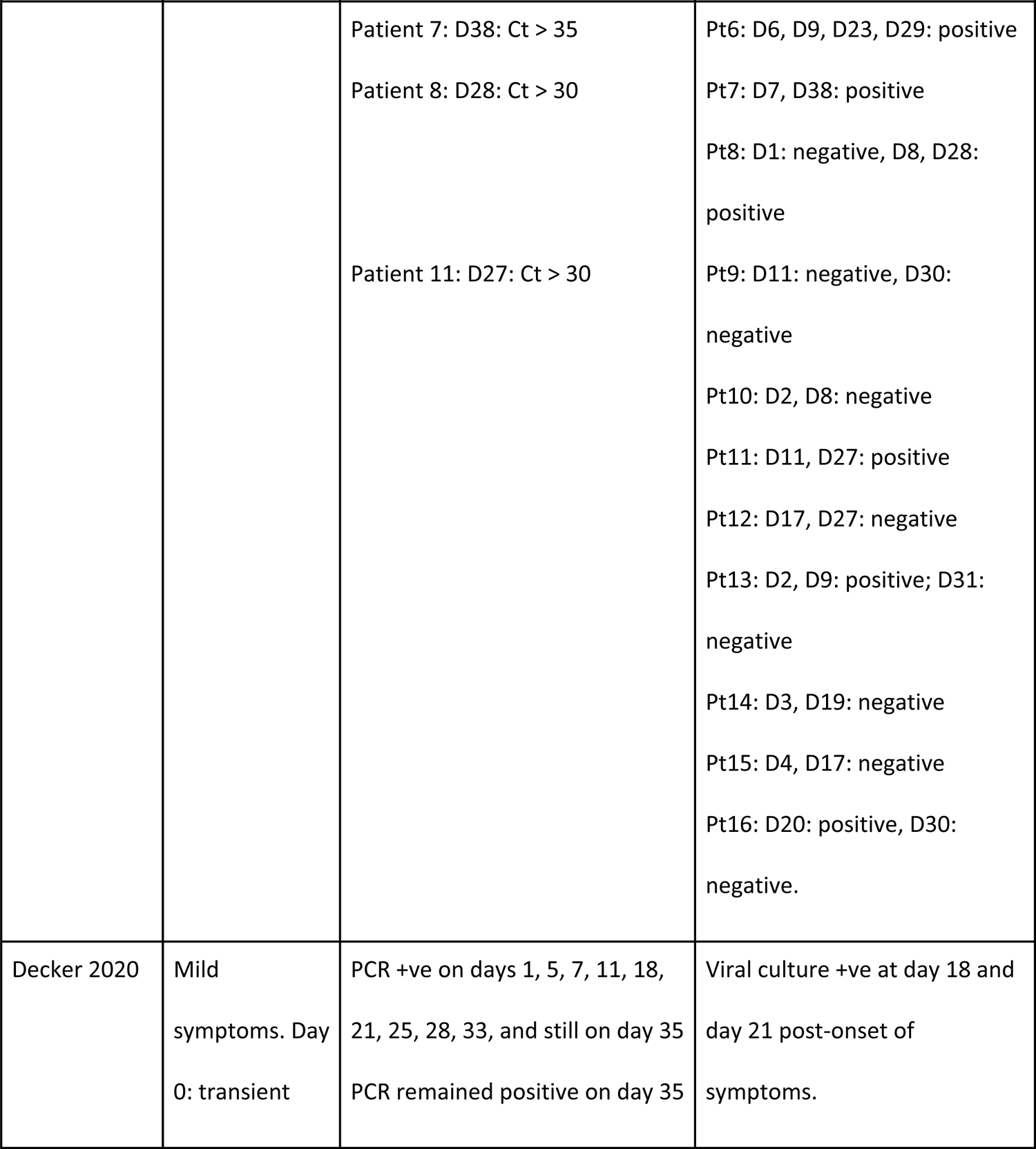
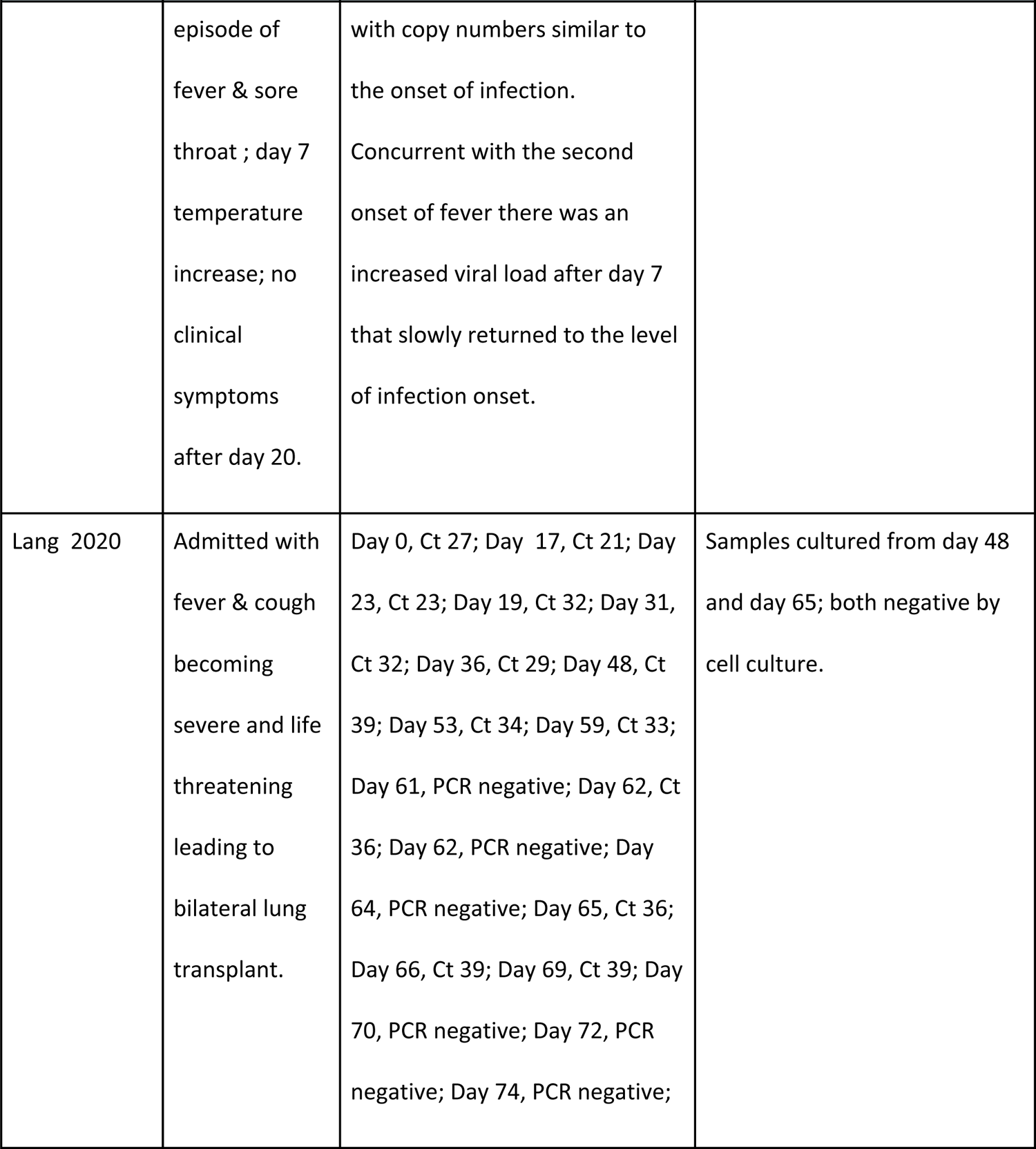
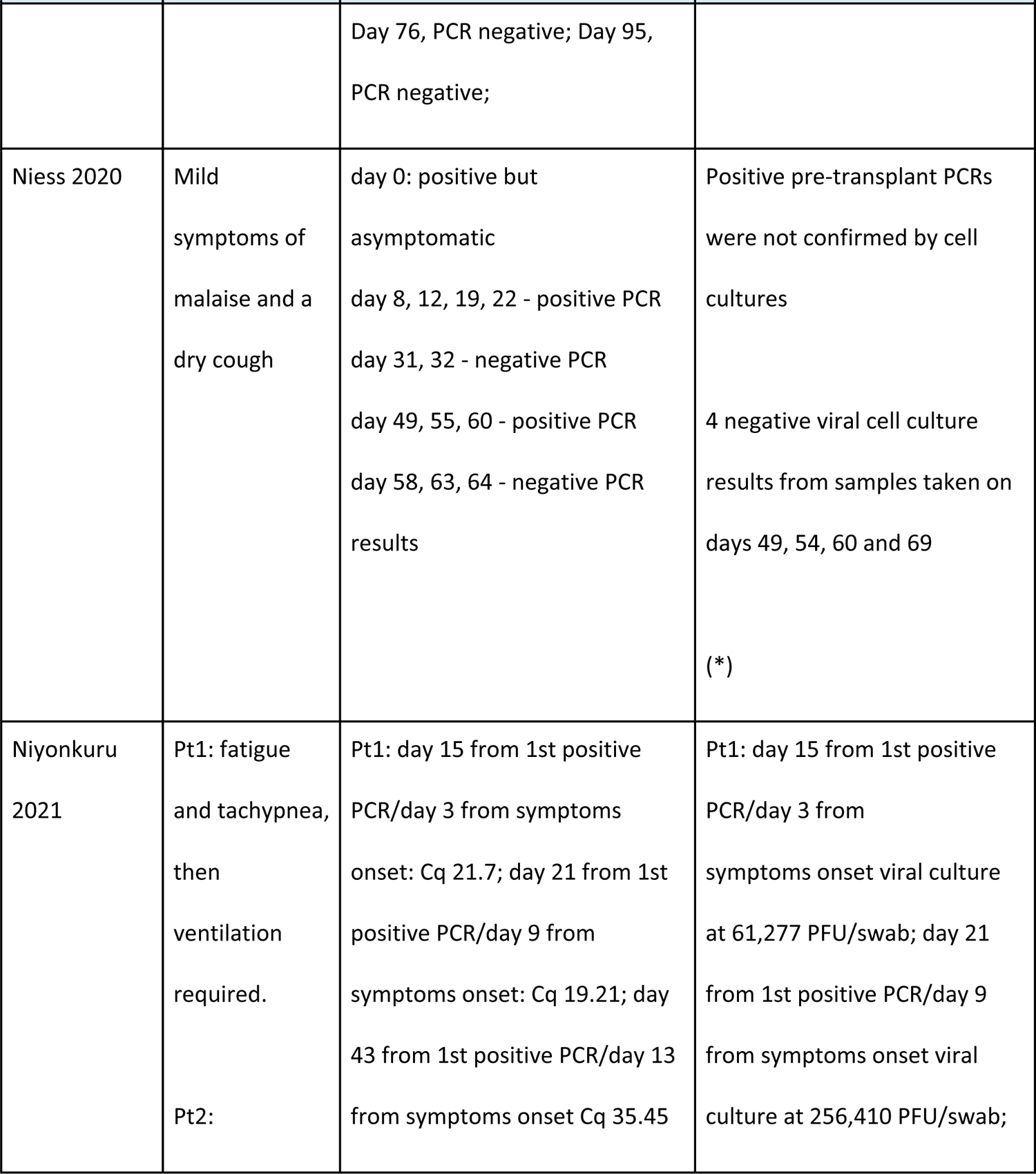
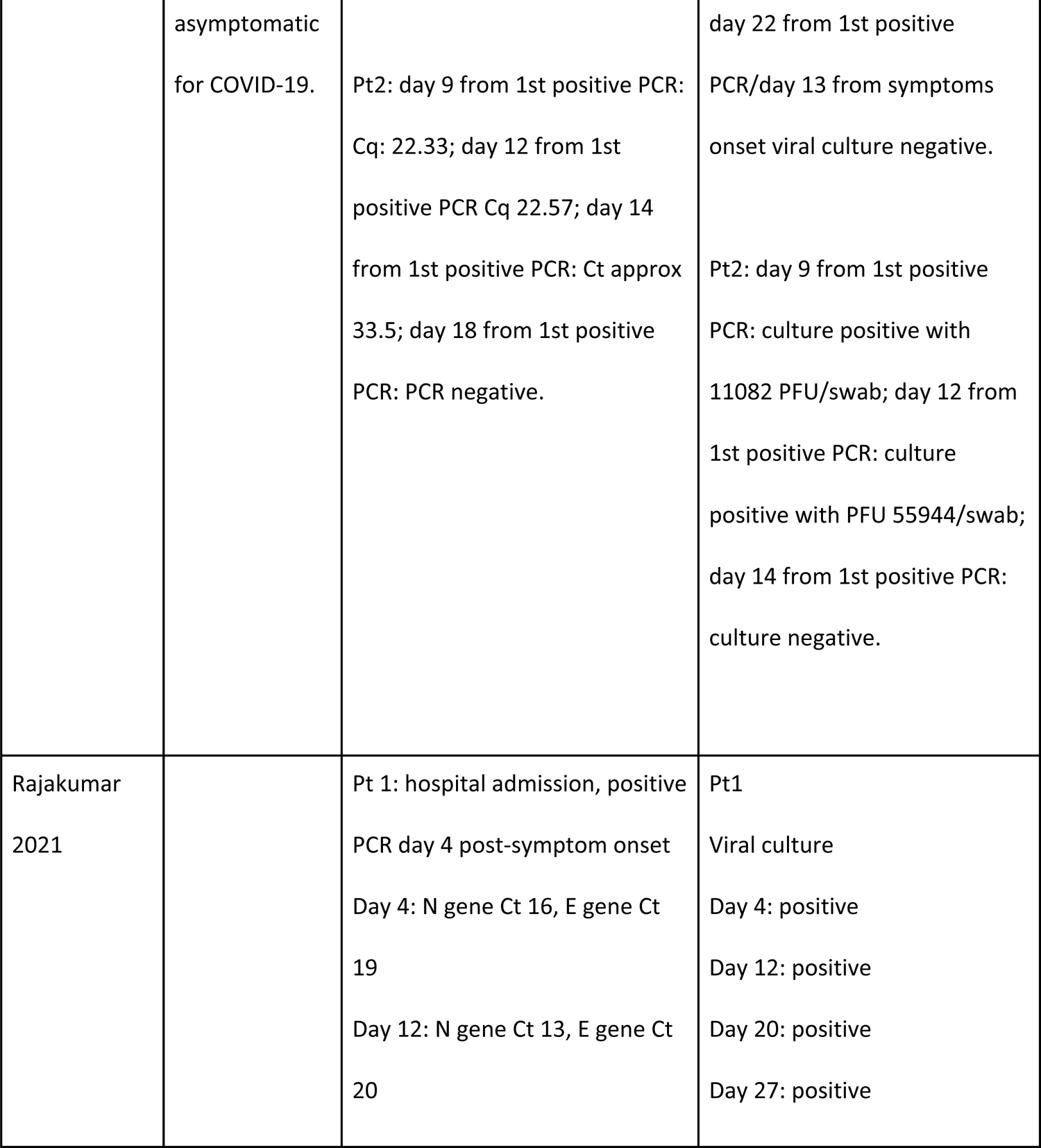
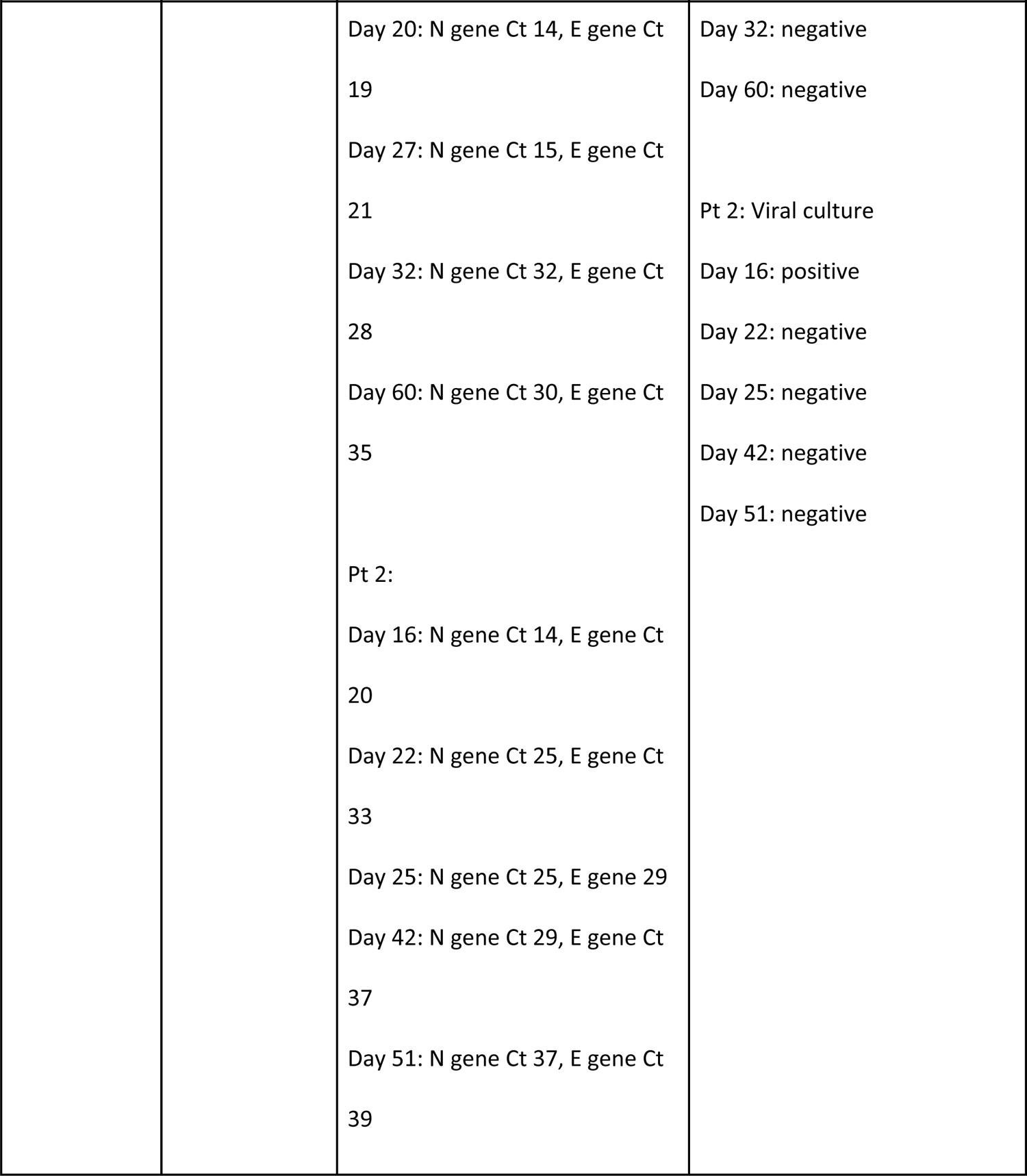
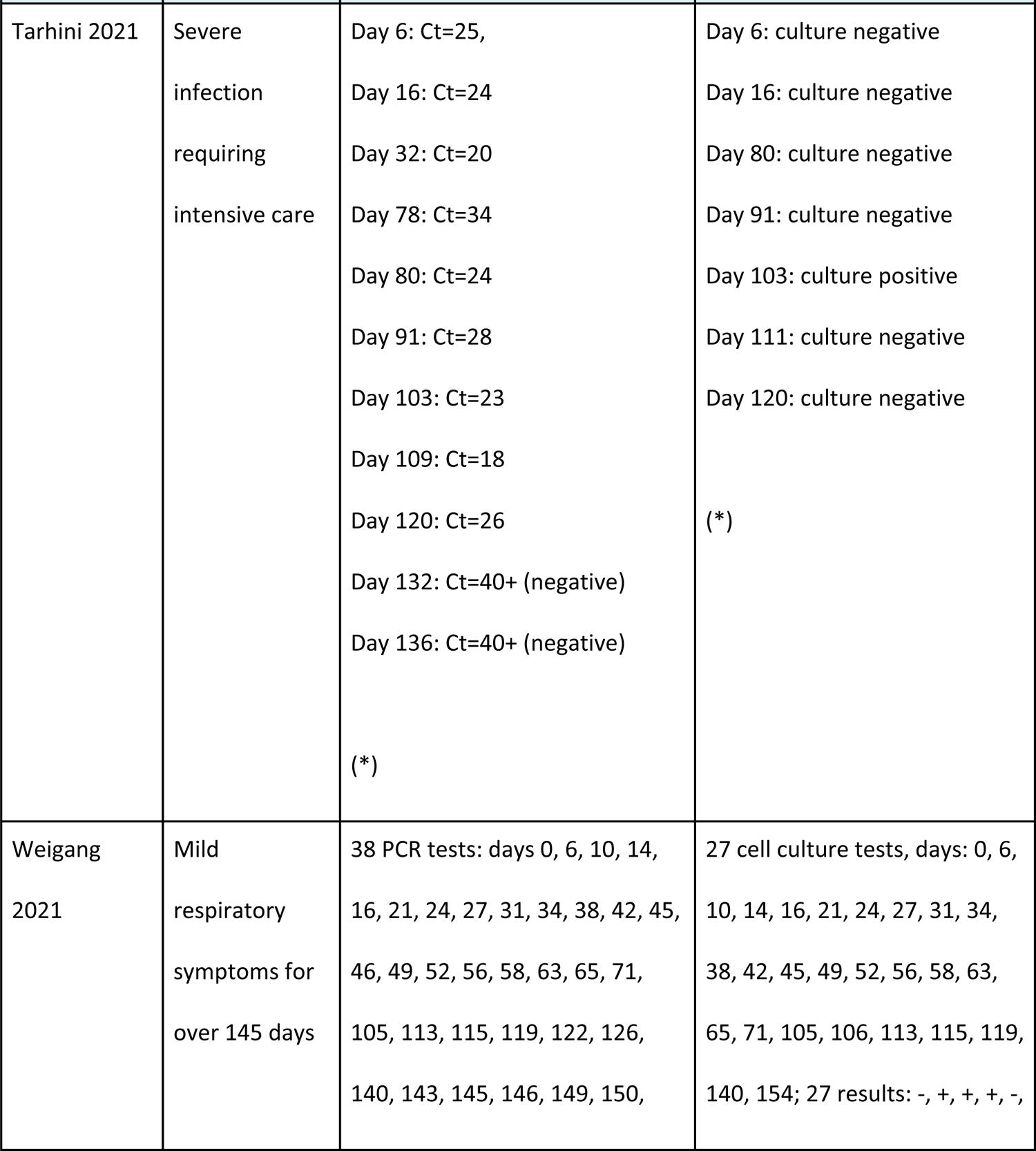
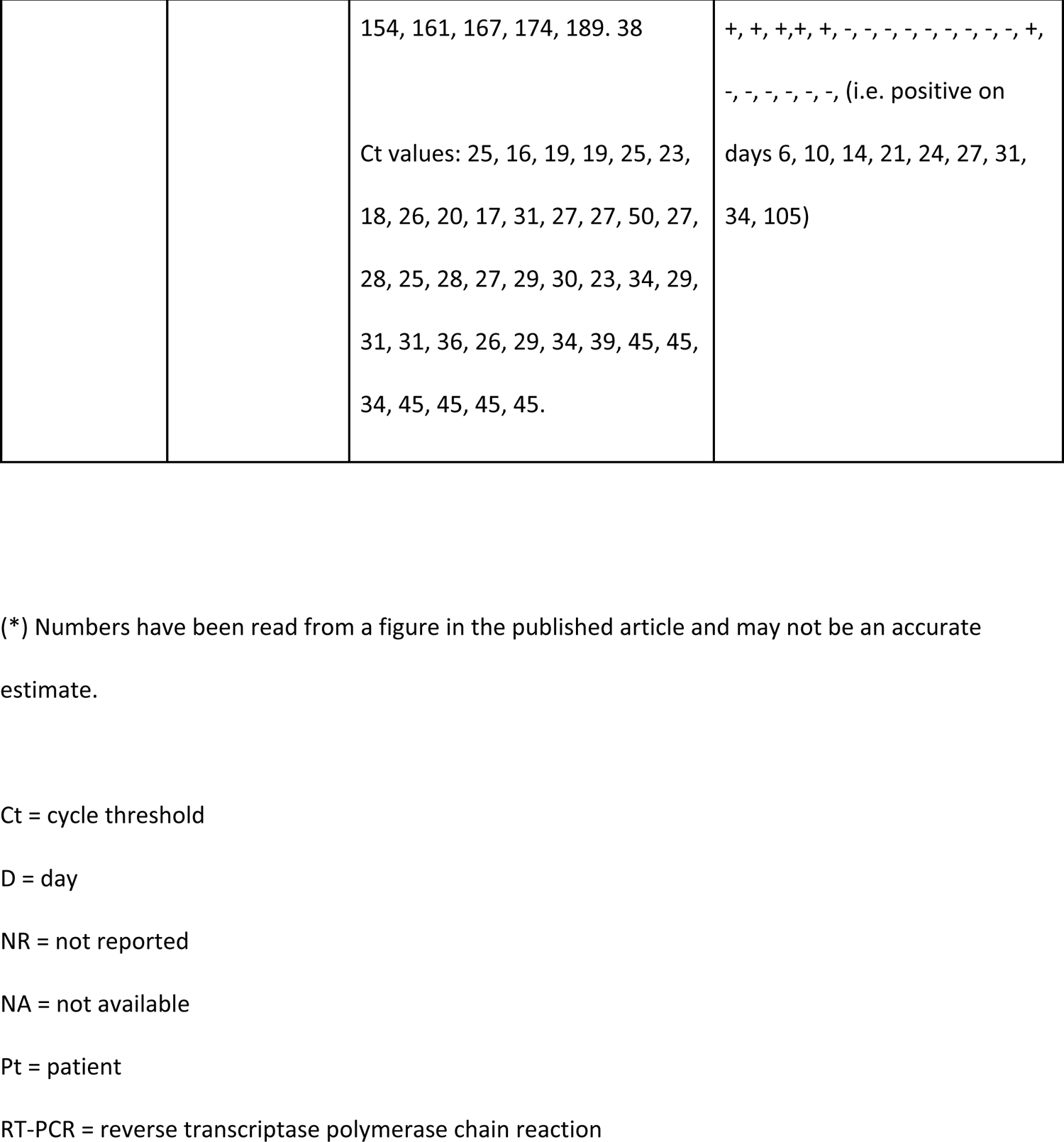
PCR cycle counts/log copies and viral culture results of included studies.

| Study ID | Symptoms,<br>days reported | RT-PCR Cycle count/log copies<br>results | Viral culture results (days) |
| --- | --- | --- | --- |
| Alshukairi<br>2021 | NR no viral<br>culture at<br>admission/ons<br>et of<br>symptoms | Pt1: D3: positive, Ct NA, D26:<br>positive, 22.87<br>Pt2: D3: positive, 11.58; D17:<br>positive, 23.12<br>Pt3: D6: positive, 8.82, D19:<br>positive, 13.88<br>Pt4: D4: positive, 10.38; D12:<br>positive, 27.57<br>Pt5: D1: positive, 2.8, D9:<br>positive, 14.84 | Pt1: D26: negative<br>Pt2: D17: negative<br>Pt3: D19: negative<br>Pt4: D12: negative<br>Pt5: D9: negative |
| Aydillo 2020<br>* | All<br>symptomatic;<br>variable<br>severity | Pt1: D3: 22.5, D27: 34<br>Pt2: D2: 18, D27: 32<br>Pt4: D5: 19, D28: 18.5, D50: 27.5<br>Pt6: D18: 21, D27: 21.5, D33:<br>30.5, D38: 20, D45: 19.5, D52: | Pt1 D3: positive, D27: negative<br>Pt2: D2: positive, D27, negative<br>Pt4: D5: positive, D28: positive,<br>D50: negative<br>Pt6: D18: negative, D27:<br>negative, D33: negative, D38: |
|  |  | 20, D62: 18.5<br>Pt8: D1: 22, D17: 21, D22: 22,<br>D37: 35<br>Pt9: D4: 13, D8: 22, D46: 33.5,<br>D56: 27, D82: 38<br>Pt13: D2: 21.5, D50: 31.5<br>Pt14: D8: 17.5 | negative, D45: positive, D52:<br>positive, D62: positive<br>Pt8: D1: positive, D17:<br>positive, D22: negative, D37:<br>negative<br>Pt9: D4: positive, D8: positive,<br>D46: negative, D56: negative,<br>D82: negative<br>Pt13: D2: positive, D50:<br>negative<br>Pt14: D8: positive |
| Benotmane<br>2021 |  | Patient 6: D23: Ct > 30; D29: Ct ><br>35 | Pt1: D5, D14: negative;<br>Pt2: D21, D29: negative;<br>Pt3: D25, D39: negative<br>Pt4: D2: positive, D9, D16, D35:<br>negative<br>Pt5: D1, D8: positive; D23, D30:<br>negative |

| Study ID | Symptoms, days reported | RT-PCR Cycle count/log copies results | Viral culture results (days) |
| --- | --- | --- | --- |
|  |  | Patient 7: D38: Ct > 35<br>Patient 8: D28: Ct > 30<br>Patient 11: D27: Ct > 30 | Pt6: D6, D9, D23, D29: positive<br>Pt7: D7, D38: positive<br>Pt8: D1: negative, D8, D28: positive<br>Pt9: D11: negative, D30: negative<br>Pt10: D2, D8: negative<br>Pt11: D11, D27: positive<br>Pt12: D17, D27: negative<br>Pt13: D2, D9: positive; D31: negative<br>Pt14: D3, D19: negative<br>Pt15: D4, D17: negative<br>Pt16: D20: positive, D30: negative. |
| Decker 2020 | Mild symptoms. Day 0: transient | PCR +ve on days 1, 5, 7, 11, 18, 21, 25, 28, 33, and still on day 35<br>PCR remained positive on day 35 | Viral culture +ve at day 18 and day 21 post-onset of symptoms. |
|  | episode of fever & sore throat ; day 7 temperature increase; no clinical symptoms after day 20. | with copy numbers similar to the onset of infection.<br>Concurrent with the second onset of fever there was an increased viral load after day 7 that slowly returned to the level of infection onset. |  |
| Lang 2020 | Admitted with fever & cough becoming severe and life threatening leading to bilateral lung transplant. | Day 0, Ct 27; Day 17, Ct 21; Day 23, Ct 23; Day 19, Ct 32; Day 31, Ct 32; Day 36, Ct 29; Day 48, Ct 39; Day 53, Ct 34; Day 59, Ct 33; Day 61, PCR negative; Day 62, Ct 36; Day 62, PCR negative; Day 64, PCR negative; Day 65, Ct 36; Day 66, Ct 39; Day 69, Ct 39; Day 70, PCR negative; Day 72, PCR negative; Day 74, PCR negative; | Samples cultured from day 48 and day 65; both negative by cell culture. |
|  |  | Day 76, PCR negative; Day 95, PCR negative; |  |
| Niess 2020 | Mild symptoms of malaise and a dry cough | day 0: positive but asymptomatic<br>day 8, 12, 19, 22 - positive PCR<br>day 31, 32 - negative PCR<br>day 49, 55, 60 - positive PCR<br>day 58, 63, 64 - negative PCR results | Positive pre-transplant PCRs were not confirmed by cell cultures<br>4 negative viral cell culture results from samples taken on days 49, 54, 60 and 69<br>(*) |
| Niyonkuru 2021 | Pt1: fatigue and tachypnea, then ventilation required.<br>Pt2: | Pt1: day 15 from 1st positive PCR/day 3 from symptoms onset: Cq 21.7; day 21 from 1st positive PCR/day 9 from symptoms onset: Cq 19.21; day 43 from 1st positive PCR/day 13 from symptoms onset Cq 35.45 | Pt1: day 15 from 1st positive PCR/day 3 from symptoms onset viral culture at 61,277 PFU/swab; day 21 from 1st positive PCR/day 9 from symptoms onset viral culture at 256,410 PFU/swab; |
|  | asymptomatic for COVID-19. | Pt2: day 9 from 1st positive PCR: Cq: 22.33; day 12 from 1st positive PCR Cq 22.57; day 14 from 1st positive PCR: Ct approx 33.5; day 18 from 1st positive PCR: PCR negative. | day 22 from 1st positive PCR/day 13 from symptoms onset viral culture negative.<br>Pt2: day 9 from 1st positive PCR: culture positive with 11082 PFU/swab; day 12 from 1st positive PCR: culture positive with PFU 55944/swab; day 14 from 1st positive PCR: culture negative. |
| Rajakumar 2021 |  | Pt 1: hospital admission, positive PCR day 4 post-symptom onset<br>Day 4: N gene Ct 16, E gene Ct 19<br>Day 12: N gene Ct 13, E gene Ct 20 | Pt1<br>Viral culture<br>Day 4: positive<br>Day 12: positive<br>Day 20: positive<br>Day 27: positive |
|  |  | Day 20: N gene Ct 14, E gene Ct 19<br>Day 27: N gene Ct 15, E gene Ct 21<br>Day 32: N gene Ct 32, E gene Ct 28<br>Day 60: N gene Ct 30, E gene Ct 35<br>Pt 2:<br>Day 16: N gene Ct 14, E gene Ct 20<br>Day 22: N gene Ct 25, E gene Ct 33<br>Day 25: N gene Ct 25, E gene 29<br>Day 42: N gene Ct 29, E gene Ct 37<br>Day 51: N gene Ct 37, E gene Ct 39 | Day 32: negative<br>Day 60: negative<br>Pt 2: Viral culture<br>Day 16: positive<br>Day 22: negative<br>Day 25: negative<br>Day 42: negative<br>Day 51: negative |

| Study ID | Symptoms, days reported | RT-PCR Cycle count/log copies results | Viral culture results (days) |
| --- | --- | --- | --- |
| Tarhini 2021 | Severe infection requiring intensive care | Day 6: Ct=25,<br>Day 16: Ct=24<br>Day 32: Ct=20<br>Day 78: Ct=34<br>Day 80: Ct=24<br>Day 91: Ct=28<br>Day 103: Ct=23<br>Day 109: Ct=18<br>Day 120: Ct=26<br>Day 132: Ct=40+ (negative)<br>Day 136: Ct=40+ (negative)<br>(*) | Day 6: culture negative<br>Day 16: culture negative<br>Day 80: culture negative<br>Day 91: culture negative<br>Day 103: culture positive<br>Day 111: culture negative<br>Day 120: culture negative<br>(*) |
| Weigang 2021 | Mild respiratory symptoms for over 145 days | 38 PCR tests: days 0, 6, 10, 14, 16, 21, 24, 27, 31, 34, 38, 42, 45, 46, 49, 52, 56, 58, 63, 65, 71, 105, 113, 115, 119, 122, 126, 140, 143, 145, 146, 149, 150, | 27 cell culture tests, days: 0, 6, 10, 14, 16, 21, 24, 27, 31, 34, 38, 42, 45, 49, 52, 56, 58, 63, 65, 71, 105, 106, 113, 115, 119, 140, 154; 27 results: -, +, +, +, -, |
|  |  | 154, 161, 167, 174, 189. 38<br>Ct values: 25, 16, 19, 19, 25, 23,<br>18, 26, 20, 17, 31, 27, 27, 50, 27,<br>28, 25, 28, 27, 29, 30, 23, 34, 29,<br>31, 31, 36, 26, 29, 34, 39, 45, 45,<br>34, 45, 45, 45, 45. | +, +, +, +, +, -, -, -, -, -, -, -, -, +,<br>-, -, -, -, -, -, (i.e. positive on<br>days 6, 10, 14, 21, 24, 27, 31,<br>34, 105) |

(\*) Numbers have been read from a figure in the published article and may not be an accurate estimate.
Ct = cycle threshold
D = day
NR = not reported
NA = not available
Pt = patient
RT-PCR = reverse transcriptase polymerase chain reaction

In response to our first study question on the correlation between proxy indicators of viral burden and infectiousness, the data from Figures 2 and 3 and Table 3 indicate a correlation between viral burden (measured as log copies or Cq/Ct) and probable infectiousness. The data suggests that earlier symptom onset is related to the likelihood of shedding replication-competent virus (Figure 2). The median time for a positive culture from onset of symptoms was 16 days (IQR 8 to 27; range 1-105, mean 19.7 days n = 59 cultures performed). The median for a negative culture was 40 days, mean 40.2 days (IQR 22 to 60; range 1-119, n = 107 cultures performed).

Five patients reported by Alshukairi et al were all culture-negative; this was in samples taken on days 9,12, 17,18 and day 26 since symptom onset, respectively ^7^.

Seven stem cell transplant recipients from the Aydillo et al letter who had repeated culture assessments fluctuated in and out of shedding replication-competent virus. One who had a single culture attempt was positive with an estimated Ct of 17.5 ^6^. Patient MSK-6 shed replication competent virus until day 62 from symptom onset with an estimated Ct of 19. MSK-6 showed the longest duration infectious transplant recipient from the Aydillo et al letter^6^. Eight kidney transplant patients described by Benotmane and colleagues had positive viral cultures ^8^.

For six patients we could identify the duration of probable infectiousness, which ranged from 8 days (patient 5: day 1 to 8) to 32 days (patient 7: day 7 to 38). Four patients were infectious with reported Cts> 30 based on the individual platforms that were used to perform the Cts.

Rajakumar et al^15^ described two cardiac transplant patients: viral culture found replication-competent virus in samples from one patient on day 16 and in samples from the other patient on day 4 and repeatedly up to day 27, after which all viral cultures were negative^15^.For each patient, viral culture was negative (i.e. no replication-competent virus observed) in samples with PCR cycle counts of over 25. Within the samples giving positive viral cultures, the PCR results showed that the cycle threshold for the N gene was lower than for the E gene by an average of 5.4 Ct values.

In the study by Niyonkuru et al, the duration of infectiousness in the two patients, as indicated by replication-competent virus, was 8 and 9 days (Figure 3) ^14^. A cardiac transplant patient described by Tarhini^9^ and colleagues tested culture-positive with a Ct of 23 on day 103; all other viral cultures were negative from samples with PCR Cts of 18 to over 40 ^9^.

Weigang et al^12^ described a kidney transplant patient who experienced three hospital admissions. During the first one (day zero to day 72), 19 RT-qPCR tests were performed, and alongside that viral culture was performed, showing 8/19 positive cultures (Ct values ranging from 15 to 25) and 11/19 negative (Ct values from 25 to 30). The patient was culture positive again on day 105 (Ct of 23). After re-admission at day 140 the patient was still RT-qPCR positive, but with viral culture negative; he was treated for 10 days (days 141-149) with remdesivir. Subsequently, negative RT-qPCR tests until day 189 and negative cultures suggested that the infection had resolved^12^.

A heart transplant patient described by Decker^10^ and co-workers had a positive viral culture on day 18 and day 21 with 6.2 and 6.5 log10 copies/ml. ^10^

Although the dataset was limited, we observed an inverse relationship (Ct/Cq) or direct relationship (log copies): the viral burden indicated by these methods correlated with infectiousness, as shown by the ability to produce replication-competent virus in culture. The presence of replication-competent virus reflects one of the highest grades of evidence supporting the capability for forward transmission of SARS-CoV-2^16, 17^.

The robustness of the correlation is difficult to assess because laboratory methods differ; it was not possible to pool the data to produce a summary cut-off value for infectiousness, due to these variations and due to varying time windows for sampling from patients (see Figure 2 and Table 3).

In response to our second research question (on the likelihood and duration of infectiousness among transplant recipients with SARS-CoV-2 infection) the data indicate that regardless of differences in laboratory practices, observed prolonged shedding of replication-competent virus is associated with alternating increases and decreases of viral burden over time, which in some cases may be up to around 100 days ^9 12^.

Figure 4 shows the relationship between cycle threshold, symptom onset (in days) and likelihood of shedding replication competent virus. In Benotmane, five results were reported with a CT of 30 or above for a positive culture. [8] However, the cell line used in this study is not demonstrated to support SARS-CoV-2 growth. In the other studies, despite a minimum of 10 different PCR platforms being used and different culture techniques, the culture results are insensitive to cycle thresholds above 30. The viral load estimates are affected by administration of courses of anti-viral treatment including remdesivir. See Figures 3a (Cts/Cqs) and 3b (log copies).

**Figure 4.**
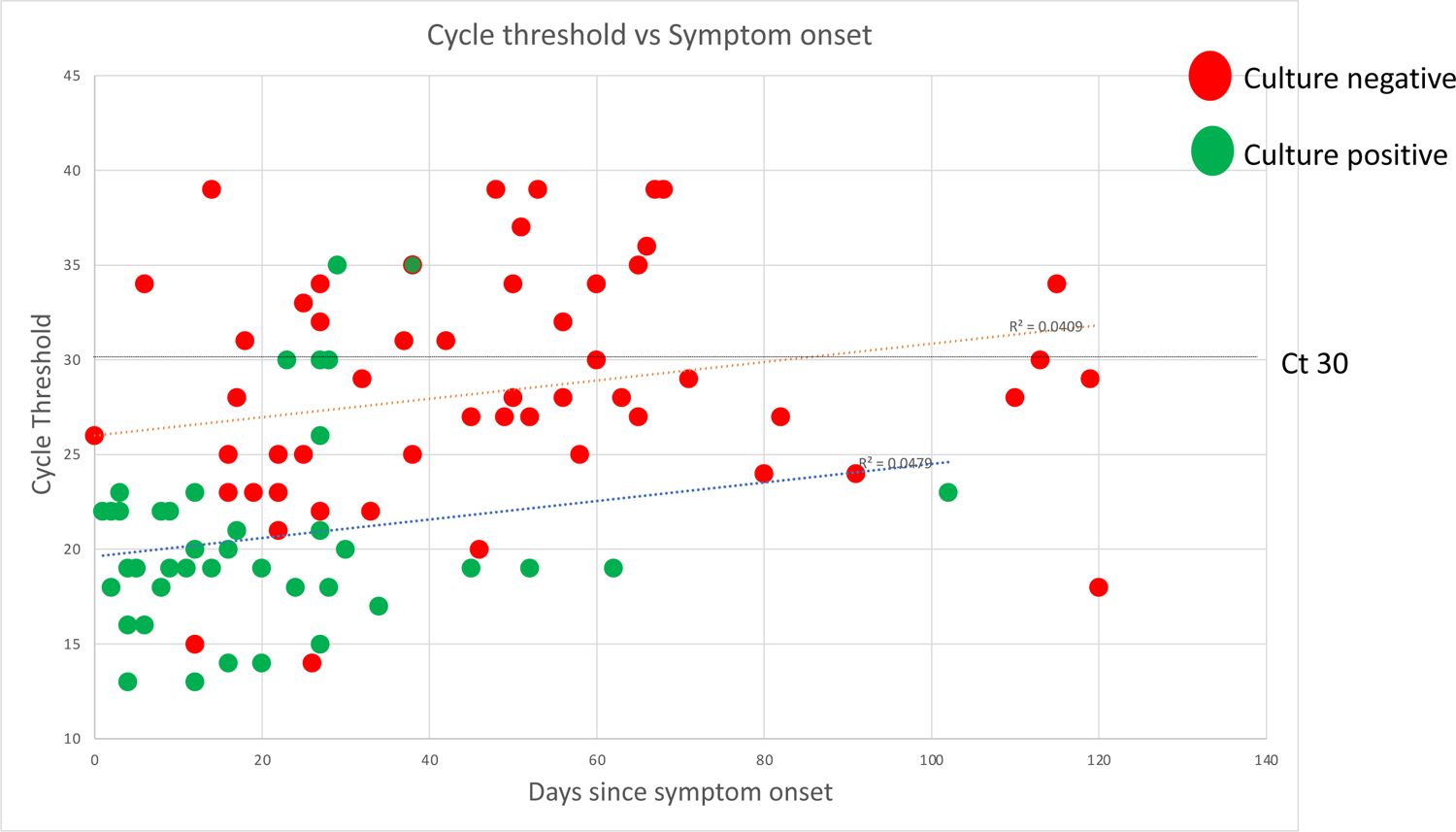
Relationship between cycle threshold, symptom onset (in days) and likelihood of shedding replication competent virus

Responding to our third research question (the influence of age, sex, underlying pathologies and degree of immunosuppression on infectiousness): at present the heterogeneity and limited amount of the available data preclude answering this question.

We are unable to answer our fourth and final research question on the relationship of vaccination status on infectiousness because no study reported on vaccination status for these transplant patients.

## Discussion

This review included 10 reports of studies using viral culture and RT-qPCR testing among 38 transplant patients with immunosuppressive treatment who experienced COVID-19 infection. The evidence indicates a relationship between indicators of viral burden (Ct, Cq or RNA log copies) and probable infectiousness as indicated by the presence of replication-competent virus. Gaps in the data remain due to variable methods and reporting and establishing summary estimates of the relationship has not been possible. The data show a long-term rise and fall of viral burden associated with the likelihood of infectiousness that in some transplant patients appears to be a sequential pattern of going in and out of infectiousness. Replication-competent virus was most commonly observed in samples with PCR Ct values under 25; one study was an exception to this by reporting viable virus at Ct>30, but the use of a cell line not typically used for SARS-CoV-2 isolation makes interpretation unclear^8^. The duration of viral RNA shedding was variable, with the longest duration reported at 105 days^12^. Our findings suggest a Ct of 30 or greater, regardless of the platform, may be used as a reasonable proxy to rule out infectious SARS-CoV-2 as there is a consistent correlation between a rising Ct value and likelihood of isolating replication-competent virus. Such a value would be useful to guide clinicians managing these difficult patients, particularly if there were repeated values in this range. Below a Ct of 30, clinicians may choose to repeat NP or throat swabs to assess the direction of the Ct values to allow a more dynamic assessment which taken in conjunction with the clinical status may faciltate decision making for isolation or antiviral treatment considerations.

Regarding our third review question on the influence of patient variables on the likelihood of the presence of infectious SARS-CoV-2, the included studies showed substantial heterogeneity; some had missing data or few cultures available, and meta-analysis or pooling was not possible. Variability in the clinical course of SARS-CoV-2 infection among transplant recipients has been reported, including observed prolonged viral shedding^18^. Antiviral drugs may impact on these observations, especially symptoms and viral burden. ^19^

Two well-designed studies on immunosuppressed patients, which we were unable to include because disaggregated data solely for transplant patients were not fully available, support our conclusions^6, 20^. While this review is limited to transplant patients, evidence suggests similar prolonged viral cultures are found in immunosuppressed cancer patients. We plan to perform a further review in this group analysing the type of cancer and the impact of immunotherapies on viral culture findings.

The transplant patient population is of particular importance: clinicians need guidance as to when to release the patient from quarantine or isolation, given the heavy burden of immunosuppression. We have tried to narrow the uncertainty and offer some general guidance as to when patients are unlikely to be shedding replication-competent virus, but clinical assessment of each patient must inform that decision because each patient and setting is different.

The strengths of this review are that we followed our published protocol, entailing rigorous literature searches, double checked data extraction and quality assessment, and a high level of clinical and epidemiological expertise input to deliberate the findings. We were also able to include data from an additional 8 transplant recipients after extensive correspondence with the study authors^6^. Limitations include the small number of studies with viral culture and serial viral load estimates among transplant patients, high variability in study design and reporting and impossibility to pool results due to the well-known variability in sensitivity across assays^21^.

Case series are conventionally considered low in the evidence hierarchy, as they may entail inherent bias in the selection of study participants and therefore have limited generalisability; however, here they are essential in providing the detailed reports needed for this unusual patient group. The case reports included here comprise some of the most detailed longitudinal reports of this patient group for whom data are needed. The evidence base is limited, however, by heterogeneous design and reporting within the studies with, for example, different observation windows for reporting of viral burden and culturability or clinical characteristics of patients.

In addition to providing appropriate care for the individual patient, ongoing transmission of SARS-CoV-2 is a concern, and immunosuppressed individuals may pose a challenge by experiencing prolonged carriage of the virus that could lead to forward transmission. Based on our findings we would offer the following general guidance to clinicians:

Physicians who are experienced with these immunosuppressed patient populations should work with public health to direct their isolation and quarantine requirements. Infectious patients with immunosuppressive treatment following solid organ or stem cell transplantation should be isolated until at least two consecutive respiratory specimens collected ≥24 hours apart demonstrate a rising RT-PCR Ct (i.e. indicating diminishing viral burden). After discharge, they should be closely followed up for SARS-CoV-2 infection for several weeks to months, depending on the individual clinical scenario.

For obtaining data, standardisation of methods is needed: each laboratory should use consistently applied platforms with suitable internal standards to calibrate the relationship between Ct and genome copy in these patient populations.

Publication of results of case series or other longitudinal study should be reported in a standardised format to avoid loss of data. We suggest observation windows should be within a short range of 3 to 7 days during the acute periods post-transplantation and during periods of rejection when higher doses of immunosuppressants are employed, depending on clinical circumstances. Each observation window should include a summary of symptoms and interventions, the reporting of PCR cycle threshold and, for samples with Ct below 30, attempts at viral culture if available. Description of patients should include past medical histories and details of treatments received. Observed drug interactions should be highlighted. Reasons for admission, discharge and changes in isolation should be clearly reported. To investigate the duration of viral shedding, studies should report the time between the first positive and the first negative viral cultures.

With additional data gathering and standardisation of methods, it will be possible for transplant physicians to develop evidence-based approaches to dealing with these patients for the benefit of the patients and their families and the community at large.

## Data Availability

All data produced in the present work are contained in the manuscript.

## Acknowledgements

We gratefully acknowledge the contributions of Drs Mini Kamboj and Jeroen van Kampen who provided additional data from their studies and helped us to progress this work.

## Author contributions

TJ, CH and JC designed the study. JB performed the literature searches. JB, TJ, SM, ER and ES, screened the studies for eligibility and performed data extraction. Additional expertise on clinical and laboratory issues was given by DE, JC, SM and ER. CH generated the data figures. All authors contributed to interpreting and writing up the results and conclusions.

## Ethics declarations

Ethics approval and consent to participate:

This is a systematic review and meta-analysis. Therefore, ethical approval is not applicable. Consent for publication:

Not applicable.

## Availability of data and materials

All data generated or analysed during this study are included in this published article [and its supplementary information files].

## Competing Interests

TJ’s competing interests are accessible at: https://restoringtrials.org/competing-interests-tom-jefferson

CJH holds grant funding from the NIHR, the NIHR School of Primary Care Research, the NIHR BRC Oxford and the World Health Organization for a series of Living rapid review on the modes of transmission of SARs-CoV-2 reference WHO registration No2020/1077093. He has received financial remuneration from an asbestos case and given legal advice on mesh and hormone pregnancy tests cases. He has received expenses and fees for his media work including occasional payments from BBC Radio 4 Inside Health and The Spectator. He receives expenses for teaching EBM and is also paid for his GP work in NHS out of hours (contract Oxford Health NHS Foundation Trust). He has also received income from the publication of a series of toolkit books and for appraising treatment recommendations in non-NHS settings. He is Director of CEBM and is an NIHR Senior Investigator.

DE holds grant funding from the Canadian Institutes for Health Research and Li Ka Shing Institute of Virology relating to the development of Covid-19 vaccines as well as the Canadian Natural Science and Engineering Research Council concerning Covid-19 aerosol transmission. He is a recipient of World Health Organization and Province of Alberta funding which supports the provision of BSL3-based SARS-CoV-2 culture services to regional investigators. He also holds public and private sector contract funding relating to the development of poxvirus-based Covid-19 vaccines, SARS-CoV-2-inactivation technologies, and serum neutralization testing.

JMC holds grants from the Canadian Institutes for Health Research on acute and primary care preparedness for COVID-19 in Alberta, Canada and was the primary local Investigator for a *Staphylococcus aureus* vaccine study funded by Pfizer for which all funding was provided only to the University of Calgary. He is co-investigator on a WHO funded study using integrated human factors and ethnography approaches to identify and scale innovative IPC guidance implementation supports in primary care with a focus on low-resource settings and using drone aerial systems to deliver medical supplies and PPE to remote First Nations communities during the COVID-19 pandemic. He also received support from the Centers for Disease Control and Prevention (CDC) to attend an Infection Control Think Tank Meeting. He is a member and Chair of the WHO Infection Prevention and Control Research and Development Expert Group for COVID-19 and a member of the WHO Health Emergencies Programme (WHE) Ad-hoc COVID-19 IPC Guidance Development Group, both of which provide multidisciplinary advice to the WHO and for which no funding is received and from which no funding recommendations are made for any WHO contracts or grants. He is also a member of the Cochrane Acute Respiratory Infections Working Group.

JB is a major shareholder in the Trip Database search engine (www.tripdatabase.com) as well as being an employee. In relation to this work Trip has worked with a large number of organisations over the years, none have any links with this work. The main current projects are with AXA and SARS-CoV-2 (WHO Registration 2020/1077093-0) and is part of the review group carrying out rapid reviews for Collateral Global. He worked on Living rapid literature review on the modes of transmission of SARS-CoV-2 and a scoping review of systematic reviews and meta-analyses of interventions designed to improve vaccination uptake (WHO Registration 2021/1138353-0).

ECR was a member of the European Federation of Neurological Societies (EFNS) / European Academy of Neurology (EAN) Scientist Panel, Subcommittee of Infectious Diseases (2013 to 2017). Since 2021, she is a member of the International Parkinson and Movement Disorder Society (MDS) Multiple System Atrophy Study Group, the Mild Cognitive Impairment in Parkinson Disease Study Group, and the Infection Related Movement Disorders Study Group. She was an External Expert and sometimes Rapporteur for COST proposals (2013, 2016, 2017, 2018, 2019) for Neurology projects. She is a Scientific Officer for the Romanian National Council for Scientific Research.

AP holds grants from the NIHR School for Primary Care Research.

IJO and EAS have no interests to disclose.

SM is a pharmacist working for the Italian National Health System since 2002 and a member of one of the three Institutional Review Boards of Emilia-Romagna Region (Comitato Etico Area Vasta Emilia Centro) since 2018.

## List of supplementary files

Supplementary file. Literature search strategy.

Supplementary file. List of excluded studies, with reasons.

## References

1. Jering, K.S., et al., Excess mortality in solid organ transplant recipients hospitalized with COVID-19: A large-scale comparison of SOT recipients hospitalized with or without COVID-19. Clin Transplant, 2022. 36(1): p. e14492. 10.1111/ctr.14492.

2. Andersen, K.M., et al., Long-term use of immunosuppressive medicines and in-hospital COVID-19 outcomes: a retrospective cohort study using data from the National COVID Cohort Collaborative. (2665-9913 (Electronic)).

3. Fernández-Ruiz, M. and J.M. Aguado, Severe acute respiratory syndrome coronavirus 2 infection in the stem cell transplant recipient - clinical spectrum and outcome. Curr Opin Infect Dis, 2021. 34(6): p. 654–662. 10.1097/qco.0000000000000790.

4. Jefferson, T., et al., Viral cultures, PCR Cycle threshold values and viral load estimation for COVID-19 infectious potential assessment in transplant patients: systematic review - Protocol Version 30 December 2021. medRxiv, 2022: p. 2021.12.30.21268509. 10.1101/2021.12.30.21268509.

5. Page, M.J., et al., The PRISMA 2020 statement: an updated guideline for reporting systematic reviews. BMJ, 2021. 372: p. n71. 10.1136/bmj.n71.

6. Aydillo, T., et al., Shedding of Viable SARS-CoV-2 after Immunosuppressive Therapy for Cancer. New England Journal of Medicine, 2020. 383(26): p. 2586–2588. 10.1056/NEJMc2031670.

7. Alshukairi, A.N., et al., Test-based de-isolation in COVID-19 immunocompromised patients: Cycle threshold value versus SARS-CoV-2 viral culture. International Journal of Infectious Diseases, 2021. 108: p. 112–115. 10.1016/j.ijid.2021.05.027.

8. Benotmane, I., et al., Long-term shedding of viable SARS-CoV-2 in kidney transplant recipients with COVID-19. American Journal of Transplantation, 2021. 21(8): p. 2871–2875. https://doi.org/10.1111/ajt.16636.

9. Tarhini, H., et al., Long-Term Severe Acute Respiratory Syndrome Coronavirus 2 (SARS-CoV-2) Infectiousness Among Three Immunocompromised Patients: From Prolonged Viral Shedding to SARS-CoV-2 Superinfection. The Journal of Infectious Diseases, 2021. 223(9): p. 1522–1527. 10.1093/infdis/jiab075.

10. Decker, A., et al., Prolonged SARS-CoV-2 shedding and mild course of COVID-19 in a patient after recent heart transplantation. Am J Transplant, 2020. 20(11): p. 3239–3245. 10.1111/ajt.16133.

11. Niess, H., et al., Liver transplantation in a patient after COVID-19 - Rapid loss of antibodies and prolonged viral RNA shedding. Am J Transplant, 2021. 21(4): p. 1629–1632. 10.1111/ajt.16349.

12. Weigang, S., et al., Within-host evolution of SARS-CoV-2 in an immunosuppressed COVID-19 patient: a source of immune escape variants. medRxiv, 2021: p. 2021.04.30.21256244. 10.1101/2021.04.30.21256244.

13. Lang, C., et al., Lung transplantation for COVID-19-associated acute respiratory distress syndrome in a PCR-positive patient. The Lancet Respiratory Medicine, 2020. 8(10): p. 1057–1060. 10.1016/S2213-2600(20)30361-1.

14. Niyonkuru, M., et al., Prolonged viral shedding of SARS-CoV-2 in two immunocompromised patients, a case report. BMC Infectious Diseases, 2021. 21(1): p. 743. 10.1186/s12879-021-06429-5.

15. Rajakumar, I.A.-O., et al., Extensive environmental contamination and prolonged severe acute respiratory coronavirus-2 (SARS CoV-2) viability in immunosuppressed recent heart transplant recipients with clinical and virologic benefit with remdesivir. (1559-6834 (Electronic)).

16. Jefferson, T., et al., Transmission of Severe Acute Respiratory Syndrome Coronavirus-2 (SARS-CoV-2) from pre and asymptomatic infected individuals. A systematic review. Clinical Microbiology and Infection, 2021. https://doi.org/10.1016/j.cmi.2021.10.015.

17. Jefferson, T.H., C.; Spencer, E.; Brassey, J.; Pluddeman, A.; Onakpoya, I.; Evans, D.; Conly, J. A Hierarchical Framework for Assessing Transmission Causality of Respiratory Viruses. PrePrints, 2021. 10.20944/preprints202104.0633.v1.

18. Marinelli, T., et al., Prospective Clinical, Virologic, and Immunologic Assessment of COVID-19 in Transplant Recipients. Transplantation, 2021. 105(10): p. 2175–2183. 10.1097/tp.0000000000003860.

19. Thornton, C.S., et al., Prolonged SARS-CoV-2 infection following rituximab treatment: clinical course and response to therapeutic interventions correlated with quantitative viral cultures and cycle threshold values. Antimicrob Resist Infect Control, 2022. 11(1): p. 28. 10.1186/s13756-022-01067-1.

20. van Kampen, J.J.A., et al., Shedding of infectious virus in hospitalized patients with coronavirus disease-2019 (COVID-19): duration and key determinants. medRxiv, 2020: p. 2020.06.08.20125310. 10.1101/2020.06.08.20125310.

21. Bruce, E.A., et al., Predicting infectivity: comparing four PCR-based assays to detect culturable SARS-CoV-2 in clinical samples. EMBO Mol Med, 2021: p. e15290. 10.15252/emmm.202115290.

