## Supplementary File. List of excluded studies for "Viral cultures, Polymerase Chain Reaction Cycle Threshold Values and Viral Load Estimation for SARS-CoV-2 Infectious Potential Assessment in Hematopoietic Stem Cell and Solid Organ Transplant Patients: A Systematic Review" (from another version of this preprint: version 1, 2022-03-02)

Supplementary data. List of studies excluded after reading the full text, with reasons

| Study reference | Reason for exclusion |
| --- | --- |
| Antar AA, Yu T, Azamfirei R et al.(2021) Host factors associated with persistent SARS-CoV-2 viral RNA in COVID-19 outpatients.  Topics in Antiviral Medicine. 29(1):68. | no transplant ppts |
| Avanzato VA, Matson MJ, Seifert SN et al. Case study: Prolonged infectious SARS-CoV-2 Shedding from an asymptomatic immunocompromised individual with cancer. Cell. 2020;183(7):1901-1912.e9. doi: 10.1016/j.cell.2020.10.049. | no transplant ppts |
| Aydillo T, Gonzalez-Reiche AS, Aslam S et al. Shedding of viable SARS-CoV-2 after immunosuppressive therapy for cancer. N Engl J Med. 2020 Dec 24;383(26):2586-2588. doi: 10.1056/NEJMc2031670. | data for transplant ppts not available * |
| Baang JH, Smith C, Mirabelli C et al. Prolonged Severe Acute Respiratory Syndrome Coronavirus 2 replication in an immunocompromised patient. J Infect Dis. 2021 Jan 4;223(1):23-27. doi: 10.1093/infdis/jiaa666. | no transplant ppts |
| Sahbudak Bal Z, Ozkul A, Bilen M, Kurugol Z, Ozkinay F. The longest infectious virus shedding in a child infected with the G614 strain of SARS-CoV-2. Pediatr Infect Dis J. 2021 Jul 1;40(7):e263-e265. doi: 10.1097/INF.0000000000003158. | no transplant ppts |
| Basheer M, Saad E, Laskar O et al. Clearance of the SARS-CoV-2 Virus in an immunocompromised patient mediated by convalescent plasma without B-Cell recovery. Int J Mol Sci. 2021 Aug 18;22(16):8902. doi: 10.3390/ijms22168902. | no transplant ppts |
| Beran A, Zink E, Mhanna M et al. Transmissibility and viral replication of SARS-COV-2 in immunocompromised patients. J Med Virol. 2021; 93(7):4156-4160. doi: 10.1002/jmv.26970. | review |
| Bhat V, Chavan P, Khattry N, Gupta S. Dynamics of viral RNA load, virus culture, seroconversion & infectivity in COVID-19 patients: Implications on isolation policy. Indian J Med Res. 2021;153(5&6):585-590. doi: 10.4103/ijmr.IJMR_3564_20. | review |
| Borges V, Isidro J, Cunha M et al. Long-term evolution of SARS-CoV-2 in an immunocompromised patient with Non-Hodgkin Lymphoma. mSphere. 2021 Aug 25;6(4):e0024421. doi: 10.1128/mSphere.00244-21. | no cell culture data |
| Casarola G, D'Abbondanza M, Curcio R et al. Efficacy of convalescent plasma therapy in immunocompromised patients with COVID-19: A case report. Clin Infect Pract. 2021 Nov;12:100096. doi: 10.1016/j.clinpr.2021.100096. | no transplant ppts |
| Ceulemans LJ, Khan M, Yoo SJ et al. Persistence of SARS-CoV-2 RNA in lung tissue after mild COVID-19. Lancet Respir Med. 2021 Aug;9(8):e78-e79. doi: 10.1016/S2213-2600(21)00240-X. | no NP PCR cycle count data |
| Charmetant X, Espi M, Benotmane I et al. Comparison of infected and vaccinated transplant recipients highlights the role of Tfh and neutralizing IgG in COVID-19 protection  medRxiv 2021.07.22.21260852; doi: https://doi.org/10.1101/2021.07.22.21260852 | no NP PCR cycle count data |
| Chen RE, Gorman MJ, Zhu DY et al. Reduced antibody activity against SARS-CoV-2 B.1.617.2 Delta virus in serum of mRNA-vaccinated patients receiving TNF-α inhibitors  medRxiv 2021.09.28.21264250; doi: https://doi.org/10.1101/2021.09.28.21264250 | no transplant ppts |
| Choi B, Choudhary MC, Regan J et al. Persistence and evolution of SARS-CoV-2 in an immunocompromised host. N Engl J Med. 2020 Dec 3;383(23):2291-2293. doi: 10.1056/NEJMc2031364. | no transplant ppts |
| Clark SA, Clark LE, Pan J et al. SARS-CoV-2 evolution in an immunocompromised host reveals shared neutralization escape mechanisms. Cell. 2021 May 13;184(10):2605-2617.e18. doi: 10.1016/j.cell.2021.03.027. | no NP PCR cycle count data |
| Dergham J, Delerce J, Bedotto M, La Scola B, Moal V. Isolation of viable SARS-CoV-2 virus from feces of an immunocompromised patient suggesting a possible fecal mode of transmission. J Clin Med. 2021 Jun 18;10(12):2696. doi: 10.3390/jcm10122696. | no NP PCR cycle count data |
| Drancourt M, Cortaredona S, Melenotte C et al. SARS-CoV-2 persistent viral shedding in the context of hydroxychloroquine-azithromycin treatment. Viruses. 2021 May 12;13(5):890. doi: 10.3390/v13050890. | no transplant ppts |
| Folgueira MD, Luczkowiak J, Lasala F, Pérez-Rivilla A, Delgado R. Prolonged SARS-CoV-2 cell culture replication in respiratory samples from patients with severe COVID-19. Clin Microbiol Infect. 2021 Jun;27(6):886-891. doi: 10.1016/j.cmi.2021.02.014. | no transplant ppts |
| Frampton D, Rampling T, Cross A et al. Genomic characteristics and clinical effect of the emergent SARS-CoV-2 B.1.1.7 lineage in London, UK: a whole-genome sequencing and hospital-based cohort study. Lancet Infect Dis. 2021 Sep;21(9):1246-1256. doi: 10.1016/S1473-3099(21)00170-5. | no cell culture data |
| Gable P, Huang JY, Gilbert SE, Bollinger S, Lyons AK, Sabour S, Surie D, Biedron C, Haney T, Beshearse E, Gregory CJ, Seely KA, Clemmons NS, Patil N, Kothari A, Gulley T, Garner K, Anderson K, Thornburg NJ, Halpin AL, McDonald LC, Kutty PK, Brown AC; Centers for Disease Control and Prevention COVID-19 Laboratory Task Force. A comparison of less invasive Severe Acute Respiratory Syndrome Coronavirus 2 (SARS-CoV-2) Diagnostic Specimens in Nursing Home Residents-Arkansas, June-August 2020. Clin Infect Dis. 2021 Jul 15;73(Suppl 1):S58-S64. doi: 10.1093/cid/ciab310. | no transplant ppts |
| Gaspar-Rodríguez A, Padilla-González A, Rivera-Toledo E. Coronavirus persistence in human respiratory tract and cell culture: An overview. Braz J Infect Dis. 2021 Sep-Oct;25(5):101632. doi: 10.1016/j.bjid.2021.101632. | review |
| Glans H, Gredmark-Russ S, Olausson M et al. Shedding of infectious SARS-CoV-2 by hospitalized COVID-19 patients in relation to serum antibody responses. BMC Infect Dis. 2021 May 27;21(1):494. doi: 10.1186/s12879-021-06202-8. | no cell culture data |
| Gniazdowski V, Paul Morris C, Wohl S et al. Repeated Coronavirus Disease 2019 molecular testing: correlation of Severe Acute Respiratory Syndrome Coronavirus 2 culture with molecular assays and cycle thresholds. Clin Infect Dis. 2021 Aug 16;73(4):e860-e869. doi: 10.1093/cid/ciaa1616. | no transplant ppts |
| Guetl K, Moazedi-Fuerst F, Rosskopf K et al. SARS-CoV-2 positive virus culture 7 weeks after onset of COVID-19 in an immunocompromised patient suffering from X chromosome-linked agammaglobulinemia. J Infect. 2021 Mar;82(3):414-451. doi: 10.1016/j.jinf.2020.10.025. | no transplant ppts |
| Han A, Rodriguez TE, Beck ET et al. Persistent SARS-CoV-2 infectivity greater than 50 days in a case series of allogeneic peripheral blood stem cell transplant recipients. Curr Probl Cancer Case Rep. 2021 Mar;3:100057. doi: 10.1016/j.cpccr.2021.100057. | no transplant ppts |
| Hensley MK, Bain WG, Jacobs J et al. Intractable Coronavirus Disease 2019 (COVID-19) and prolonged Severe Acute Respiratory Syndrome Coronavirus 2 (SARS-CoV-2) replication in a chimeric antigen receptor-modified T-Cell therapy recipient: a case study. Clin Infect Dis. 2021 Aug 2;73(3):e815-e821. doi: 10.1093/cid/ciab072. | no transplant ppts |
| Kemp SA, Collier DA, Datir RP et al. SARS-CoV-2 evolution during treatment of chronic infection. Nature. 2021 Apr;592(7853):277-282. doi: 10.1038/s41586-021-03291-y. | no transplant ppts |
| Khatamzas E, Rehn A, Muenchhoff M et al. Emergence of multiple SARS-CoV-2 mutations in an immunocompromised host. medRxiv 2021.01.10.20248871; doi: https://doi.org/10.1101/2021.01.10.20248871 | no transplant ppts |
| Klein J, Brito AF, Trubin P et al. Longitudinal immune profiling of a SARS-CoV-2 reinfection in a solid organ transplant recipient. J Infect Dis. 2021 Oct 29:jiab553. doi: 10.1093/infdis/jiab553. | no cell culture data |
| Kociolek LK, Muller WJ, Yee R et al. Comparison of upper respiratory viral load distributions in asymptomatic and symptomatic children diagnosed with SARS-CoV-2 infection in pediatric hspital testing programs. J Clin Microbiol. 2020 Dec 17;59(1):e02593-20. doi: 10.1128/JCM.02593-20. | no cell culture data |
| Koff AG, Laurent-Rolle M, Hsu JC, Malinis M. Prolonged incubation of severe acute respiratory syndrome coronavirus 2 (SARS-CoV-2) in a patient on rituximab therapy. Infect Control Hosp Epidemiol. 2021 Oct;42(10):1286-1288. doi: 10.1017/ice.2020.1239. | no transplant ppts |
| Lacson E Jr, Weiner D, Majchrzak K et al. Prolonged live SARS-CoV-2 shedding in a maintenance dialysis patient. Kidney Med. 2021 Mar-Apr;3(2):309-311. doi: 10.1016/j.xkme.2020.12.001. | no NP PCR cycle count data |
| Leitão IC, Calil PT, Galliez RM et al. Prolonged SARS-CoV-2 positivity in immunocompetent patients: virus isolation, genomic integrity, and transmission risk. Microbiol Spectr. 2021 Dec 22;9(3):e0085521. doi: 10.1128/Spectrum.00855-21. | no transplant ppts |
| Leung WF, Chorlton S, Tyson J et al. COVID-19 in an immunocompromised host: persistent shedding of viable SARS-CoV-2 and emergence of multiple mutations: a case report. Int J Infect Dis. 2022 Jan;114:178-182. doi: 10.1016/j.ijid.2021.10.045. | no transplant ppts |
| Mendes-Correa MC, Ghilardi F, Salomão MC et al. SARS-CoV-2 shedding, infectivity and evolution in an immunocompromised adult patient. medRxiv 2021.06.11.21257717; doi: https://doi.org/10.1101/2021.06.11.21257717 | no transplant ppts |
| Monrad I, Sahlertz SR, Nielsen SSF et al. Persistent Severe Acute Respiratory Syndrome Coronavirus 2 infection in immunocompromised host displaying treatment induced viral evolution. Open Forum Infect Dis. 2021 Jun 4;8(7):ofab295. doi: 10.1093/ofid/ofab295. | no transplant ppts |
| Nemudryi A, Nemudraia A, Wiegand T et al. SARS-CoV-2 genomic surveillance identifies naturally occurring truncation of ORF7a that limits immune suppression. Cell Rep. 2021 Jun 1;35(9):109197. doi: 10.1016/j.celrep.2021.109197. | no cell culture data |
| Nomura T, Kitagawa H, Omori K et al. Duration of infectious virus shedding in patients with severe coronavirus disease 2019 who required mechanical ventilation. J Infect Chemother. 2022 Jan;28(1):19-23. doi: 10.1016/j.jiac.2021.09.006. | no transplant ppts |
| Pérez-Lago L, Aldámiz-Echevarría T, García-Martínez R et al. Different within-host viral evolution dynamics in severely immunosuppressed cases with persistent SARS-CoV-2. Biomedicines. 2021 Jul 13;9(7):808. doi: 10.3390/biomedicines9070808. | no transplant ppts |
| Purpura LJ, Chang M, Annavajhala MK et al. Prolonged severe acute respiratory syndrome coronavirus 2 persistence, attenuated immunologic response, and viral evolution in a solid organ transplant patient. Am J Transplant. 2021 Sep 12. doi: 10.1111/ajt.16837. | no cell culture data |
| Rhee C, Kanjilal S, Baker M, Klompas M. Duration of Severe Acute Respiratory Syndrome Coronavirus 2 (SARS-CoV-2) infectivity: when is it safe to discontinue isolation? Clin Infect Dis. 2021 Apr 26;72(8):1467-1474. doi: 10.1093/cid/ciaa1249. | review |
| Rodríguez-Grande C, Adán-Jiménez J, Catalán P et al. Inference of active viral teplication in cases with sustained positive reverse transcription-PCR Results for SARS-CoV-2. J Clin Microbiol. 2021 Jan 21;59(2):e02277-20. doi: 10.1128/JCM.02277-20. | no cell culture data |
| Roedl K, Heidenreich S, Pfefferle S et al. Viral dynamics of SARS-CoV-2 in critically ill allogeneic hematopoietic stem cell transplant recipients and immunocompetent patients with COVID-19. Am J Respir Crit Care Med. 2021 Jan 15;203(2):242-245. doi: 10.1164/rccm.202009-3386LE. | no NP PCR cycle count data |
| Gilad R, Oran E, Itai G et al. Evaluation of the relationship between quantitative PCR results and cell culturing of SARS2-CoV with respect to symptoms onset and Viral load – a systematic review. doi: https://doi.org/10.1101/2021.08.23.21262162 | review |
| Sepulcri C, Dentone C, Mikulska M et al. The longest persistence of viable SARS-CoV-2 with recurrence of viremia and relapsing symptomatic COVID-19 in an immunocompromised patient-a case study. Open Forum Infect Dis. 2021 Apr 28;8(11):ofab217. doi: 10.1093/ofid/ofab217. | no transplant ppts |
| Singanayagam A, Patel M, Charlett A et al. Duration of infectiousness and correlation with RT-PCR cycle threshold values in cases of COVID-19, England, January to May 2020. Euro Surveill. 2020 Aug;25(32):2001483. doi: 10.2807/1560-7917.ES.2020.25.32.2001483. | no transplant ppts |
| Surie D, Huang JY, Brown AC et al. Infectious period of Severe Acute Respiratory Syndrome Coronavirus 2 in 17 nursing home residents-Arkansas, June-August 2020. Open Forum Infect Dis. 2021 Jan 30;8(3):ofab048. doi: 10.1093/ofid/ofab048. | no transplant ppts |
| Truong TT, Ryutov A, Pandey U et al. Increased viral variants in children and young adults with impaired humoral immunity and persistent SARS-CoV-2 infection: A consecutive case series. EBioMedicine. 2021 May;67:103355. doi: 10.1016/j.ebiom.2021.103355. | no transplant ppts |
| Van Cleemput J, van Snippenberg W, Lambrechts L et al. Organ-specific genome diversity of replication-competent SARS-CoV-2. Nat Commun. 2021 Nov 16;12(1):6612. doi: 10.1038/s41467-021-26884-7. | no NP PCR cycle count data |
| van Kampen JJA, van de Vijver DAMC, Fraaij PLA et al. Duration and key determinants of infectious virus shedding in hospitalized patients with coronavirus disease-2019 (COVID-19). Nat Commun. 2021 Jan 11;12(1):267. doi: 10.1038/s41467-020-20568-4. | data for transplant ppts not available * |
| Viana LA, Cristelli MP, Ficher KN et al. Kidney transplantation in patients With SARS-CoV-2 infection: a case series report. Transplantation. 2021 Jan 1;105(1):e1-e3. doi: 10.1097/TP.0000000000003521. | no NP PCR cycle count data |
| Viratam MD, Villanueva, M, Shenoi, S et al. SARS-CoV-2 viral dynamics for symptomatic people living with HIV requiring hospitalization for COVID-19. Open Forum Infectious Diseases, Volume 7, Issue Supplement_1, October 2020, Page S338, https://doi.org/10.1093/ofid/ofaa439.737 | no cell culture data |
| Walsh KA, Spillane S, Comber L et al. The duration of infectiousness of individuals infected with SARS-CoV-2. J Infect. 2020 Dec;81(6):847-856. doi: 10.1016/j.jinf.2020.10.009. | review |
| Williamson MK, Hamilton F, Hutchings S et al. Chronic SARS-CoV-2 infection and viral evolution in a hypogammaglobulinaemic individual.  medRxiv 2021.05.31.21257591; doi: https://doi.org/10.1101/2021.05.31.21257591 | no transplant ppts |

*We contacted the corresponding authors who kindly tried to determine if we could be provided with the disaggregated data for the transplant patients, but due to data protection issues this was not possible. We are grateful to Dr Mina Kamoboj and Dr Jeroen van Kampen for their help with this.
