## Supplementary file for "Viral cultures, Polymerase Chain Reaction Cycle Threshold Values and Viral Load Estimation for SARS-CoV-2 Infectious Potential Assessment in Hematopoietic Stem Cell and Solid Organ Transplant Patients: A Systematic Review" (from another version of this preprint: version 1, 2022-03-02)

Literature search strategy and flow of studies through the screening process

Search terms

(coronavirus OR covid-19 OR SARS-CoV-2) AND (immunosuppressed OR immunocompromised OR transplant OR immunosuppression OR "immune deficient" OR HIV) AND (CPE OR “cytopathic effect” OR “Viral culture” OR “virus culture” OR vero OR "virus replication" OR "viral replication" OR "cell culture" or "viral load" OR "viral threshold" OR "log copies" OR "cycle threshold")

- **WHO Covid-19 Database** (<https://search.bvsalud.org/global-literature-on-novel-coronavirus-2019-ncov/>)

The global literature cited in the WHO COVID-19 database is updated daily (Monday through Friday) from searches of bibliographic databases, hand searching, and the addition of other expert-referred scientific articles.

- **LitCovid** (<https://www.ncbi.nlm.nih.gov/research/coronavirus/>)

A curated literature hub for tracking up-to-date scientific information about the 2019 novel Coronavirus. It is a comprehensive resource on the subject, providing central access to relevant articles in PubMed.

- **medRxiv** (<https://www.medrxiv.org/>)

A free online archive and distribution server for complete but unpublished manuscripts (preprints) in the medical, clinical, and related health sciences.

- **Google Scholar** (<https://scholar.google.com/>)

Provides a broad search for scholarly literature across many disciplines and sources: articles, theses, books, abstracts, from academic publishers, professional societies, online repositories, universities and other websites.

Search terms covered three domains to identify:

- Covid-19 papers (not needed on LitCovid and the WHO Covid-19 databases as these are specialist Covid-19 databases).
- Papers dealing with immunosuppressed populations e.g. variants of *immunosuppressed* or *immunosuppression* and specific populations such as *transplant* and *HIV*.
- Papers that explore viral culture and cycle thresholds.

We then screened within these studies to identify those including patients with transplants.
